# Predicting Cognitive Functioning in ADHD Using Population-Based MRI Across Large and Small Samples

**DOI:** 10.1101/2025.07.17.25331721

**Authors:** Farzane Lal Khakpoor, William van der Vliet, Alina Tetereva, Yue Wang, Narun Pat

**Affiliations:** Department of Psychology, University of Otago, Dunedin, New Zealand; New Zealand Brain Research Institute, Christchurch, New Zealand

**Keywords:** ADHD, Cognitive prediction, Generalizability, MRI, Neuroimaging

## Abstract

Machine learning offers a promising framework for transforming neuroimaging into predictive tools for psychiatric phenotypes. However, a major translational challenge is whether models trained on large, heterogeneous population cohorts can generalize to clinical populations without dataset-specific tuning. Here, we tested the generalizability of neuroimaging-based cognitive prediction models to children with attention-deficit/hyperactivity disorder (ADHD). Using baseline data from the Adolescent Brain Cognitive Development (ABCD) Study (*n* = 11,747), we trained prediction models across 81 MRI-derived feature sets spanning task-based fMRI contrasts, functional connectivity, structural MRI, and diffusion measures and their multimodal stacking combinations. We tested performance separately in non-ADHD participants and across increasingly stringent ADHD tiers, and further evaluated external generalization in an independent ADHD cohort (Lytle et al., 2020; *n*=79) without retraining. Internally, the stacked model integrating all feature sets achieved the highest predictive accuracy (*r*=.57) and reproduced observed group-level cognitive differences, while maintaining comparable performance across ADHD and non-ADHD groups. Externally, stacked working-memory task models generalized to the independent ADHD cohort (*r*=.44). Together, these findings provide evidence that population-trained, multimodal neuroimaging models can capture cognition-related neural variation across ADHD and non-ADHD groups, while the external validation provides preliminary evidence of transferability across cohorts and measurement contexts.

---

Machine learning holds promise for exploring brain-behaviour relationships and transforming neuroimaging into a predictive tool for psychiatric phenotypes (Sui et al., 2020). Large, population-based datasets have enabled promising advances in predicting cognitive functioning from neuroimaging (Finn et al., 2015; Sripada et al., 2020; Vieira et al., 2022). Particularly, those using multimodal neuroimaging, which integrates information from various neuroimaging types, tend to achieve relatively higher predictive performance than single modalities (Marek et al., 2022; Rasero et al., 2021; Rosenberg et al., 2018; Tetereva et al., 2022, 2025). While such datasets are ideal for training such models due to their size and diversity. They often include a mix of typically developing individuals and those with subclinical or undiagnosed conditions, which may not fully represent the neurocognitive profiles of clinical populations. Conversely, smaller clinical datasets offer diagnostic specificity but lack the statistical power for robust model training (Marek et al., 2022; Pat et al., 2022; Poldrack et al., 2020; Sui et al., 2020; Tetereva et al., 2022, 2025; Varoquaux, 2018; Wang et al., 2025).

A critical challenge in human brain mapping is determining whether predictive models trained on large, heterogeneous cohorts can directly generalize to clinical populations without further retraining or tuning. This direct generalizability is essential for translating neuroimaging into clinically useful tools and for testing the universality of brain–cognition relationships across the neurodevelopmental spectrum. Rigorous validation, both internal (within the same dataset) and external (across independent datasets), is essential to bridge this gap.

Cognitive functioning, encompassing attention, inhibition, working/declarative memory, and language, is a key psychiatric phenotype that predicts daily functioning and treatment outcomes (Abramovitch et al., 2021). Frameworks like the Research Domain Criteria (RDoC) propose that cognitive functioning varies along a transdiagnostic spectrum, spanning from typical to atypical performance, reflecting a continuum of neural and behavioral characteristics across both healthy and clinical populations (Chopra et al., 2024; Morris & Cuthbert, 2012). Disruptions in cognitive functioning can significantly impair quality of life, particularly in neurodevelopmental conditions such as attention-deficit/hyperactivity disorder (ADHD), a highly prevalent neurodevelopmental condition (Sacco et al., 2024) with deficits in several cognitive domains (Cohen-Gilbert et al., 2022; Duff & Sulla, 2015; Kofler et al., 2019; Senkowski et al., 2024), and well-documented structural and functional brain alterations (Cortese et al., 2021; Luders et al., 2009; Mooney et al., 2024; Norman et al., 2016; Seidman et al., 2005), including reduced cortical thickness in prefrontal regions, altered connectivity in attention networks, diminished activity in prefrontal and parietal regions during cognitive tasks and atypical white matter organization (Dickstein et al., 2006; Emond et al., 2009; Hoogman et al., 2019; Samea et al., 2019). This behavioral and neural pattern makes ADHD an ideal test case to assess generalisability of neuroimaging-based cognitive prediction models across populations. By examining model performance in ADHD, we can determine if identified neural patterns of cognition are robust across typical and atypical brain development (Chopra et al., 2024; Morris & Cuthbert, 2012), thereby advancing our understanding of human brain function in capturing the transdiagnostic spectrum of cognitive functioning.

To explore this, we trained cognitive prediction models using all the available neuroimaging data, quantified into 81 sets of neuroimaging features, from population-based ABCD (Jernigan et al., 2024) children and tested their performance on children with or without ADHD. Next, to assess whether cognitive prediction models trained in a large population-based dataset could transfer to a smaller clinical-focused dataset, we tested their performance using both internal validation (i.e., on children with ADHD within the ABCD) and external validation (i.e., on children with ADHD from a small-scale study (Lytle et al., 2020)). Given the ADHD-related alterations in brain morphology and functions (Cortese et al., 2021; Luders et al., 2009; Mooney et al., 2024; Norman et al., 2016; Seidman et al., 2005), some neuroimaging types may lead to more generalizable models than others. To our knowledge, this study provides the first comprehensive benchmark of model generalizability across 81 MRI-derived feature sets. We compare models trained on individual feature sets (e.g., cortical thickness measures or task-fMRI contrasts; hereafter, *single models*) with models that integrate information across feature sets using a machine-learning stacking approach (Engemann et al., 2020; Tetereva et al., 2022) (hereafter, *stacked models*). Because this was an exploratory benchmark, we did not prespecify which modalities were key or redundant; instead, we evaluated a broad feature space to identify patterns that can guide more focused future studies. These findings may guide more efficient neuroimaging data collection and analysis strategies by identifying modalities and combinations that are informative for cognitive prediction. Specifically, we tested the following hypotheses: 1) Group differences: Observed cognitive functioning scores will be lower in children with ADHD than in non-ADHD peers, and these differences should be reflected in model predictions. 2) Consistency of predictive performance: The accuracy of cognitive prediction models should not differ significantly between ADHD and non-ADHD groups. 3) Benefit of multimodal integration: Models that integrate multiple neuroimaging modalities (stacked models) should yield higher predictive accuracy in ADHD groups compared to single-modality models. 4) External validation: Models trained on large, population-based data should generalize to a smaller, independent ADHD dataset without the need for tuning, achieving comparable performance across ADHD and non-ADHD participants.

## MATERIALS AND METHODS

Figure 1 shows the overall design and information about participants from the Adolescent Brain Cognitive Development (ABCD) Study (baseline from release 5.1, N = 11,878, F = 5680, age range = 9–10) (Jernigan et al., 2024) and Lytle and colleagues’ study (N= 79, F = 14, age range = 8-10) (Lytle et al., 2020). We used the population-based ABCD to a) train the cognitive prediction models and b) internally validate the models’ performance on out-of-sample participants with ADHD within the same dataset (but not part of the model-building process). We used Lytle and colleagues’ small-scale study on ADHD (Lytle et al., 2020) to externally validate the models trained from the ABCD. Institutional Review Boards at each data collection site approved the study protocols (Clark et al., 2018; Lytle et al., 2020).

**Figure 1.**
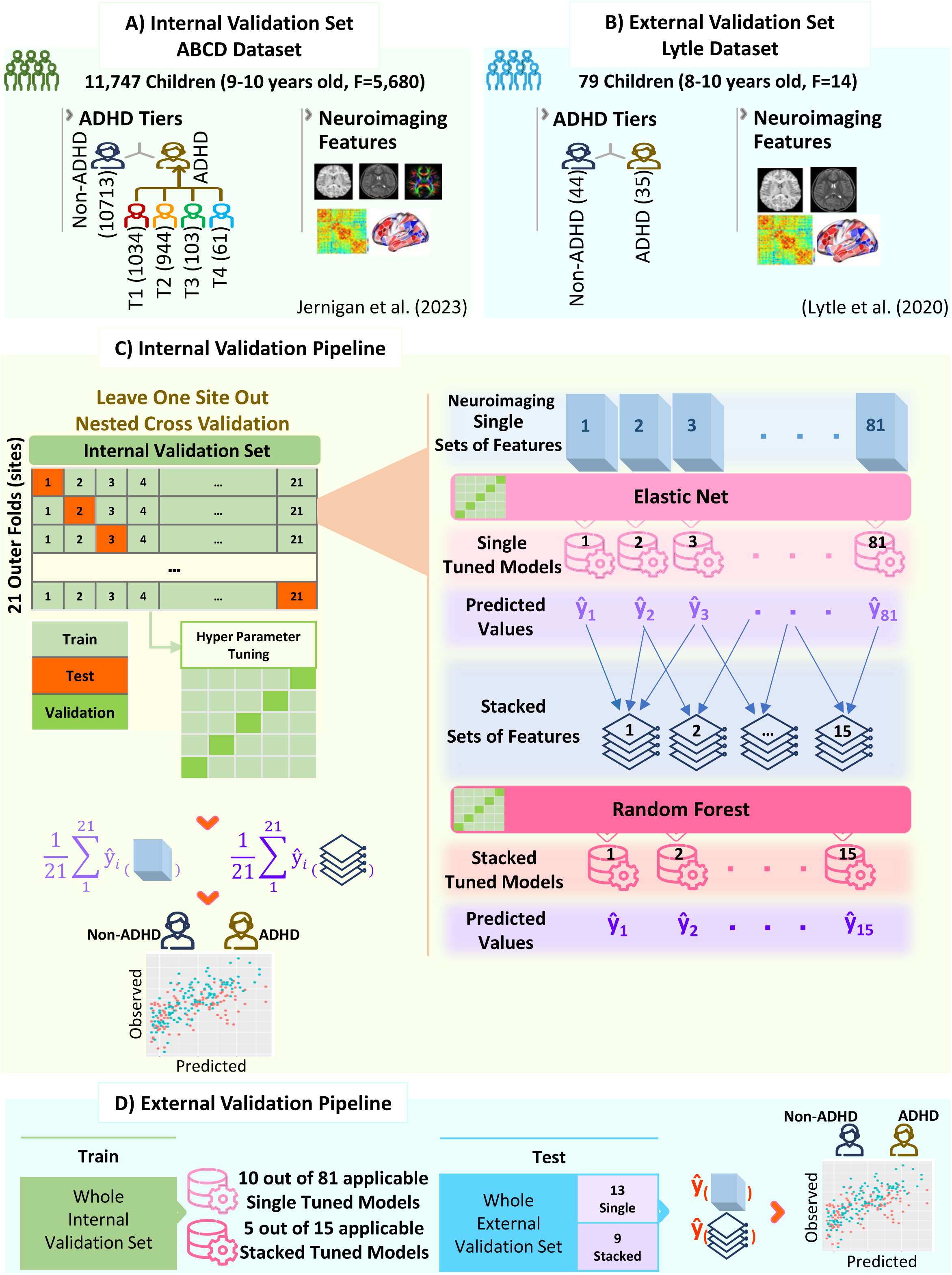
Overall design and information about participants. **(A)** and **(B)** indicate the data we extracted from the ABCD (i.e., internal validation dataset) and Lytle and colleagues’ (i.e., external validation dataset). For the tier definitions of ADHD in the ABCD, we followed Cordova and colleagues (Cordova et al., 2022). Specifically, ADHD Tier 1 included current ADHD based on parent-reported Kiddie Schedule for Affective Disorders and Schizophrenia – Computerized Version (K-SADS-COMP) (Kaufman et al., 1997), Tier 2 excluded cases with bipolar disorder, schizophrenia spectrum, or intellectual disability, Tier 3 filtered based on the teacher-rated Brief Problem Monitoring scale (BPM) (Achenbach & Rescorla, 2001) to confirm symptoms in multiple settings, and Tier 4 applied the clinical cut-off from the Child Behaviour Checklist (CBCL) attention or ADHD DSM-5 scale (Achenbach & Ruffle, 2000). **(C)** summarizes the internal validation pipeline, including the nested leave-one-site-out procedure and stacked-model construction. **(D)** shows our pipeline for externally validating the predictive performance of our models built from the ABCD. Note that not all sets of neuroimaging features provided by the ABCD are available in Lytle and colleagues’ study. As a result, when conducting external validation, we could only apply the models trained from the ABCD to 13 single and nine stacked models, as opposed to 81 single and 15 stacked models used in our internal validation.

### ADHD identification

For the ABCD, we identified ADHD using Cordova and colleagues’ (Cordova et al., 2022) tier system which approximated the Diagnostic and Statistical Manual of Mental Illnesses (DSM-5) (American Psychiatric Association, 2022) standards and progressed from least to most stringent (see Figure 1A and Supplementary Methods). Briefly, we used parent-and teacher-reported responses and ruled out comorbid conditions. Tier 1 included participants with a current ADHD diagnosis based on parent-reported responses from the K-SADS-COMP (Kaufman et al., 1997). Tier 2 refined this group by excluding individuals with bipolar disorder, schizophrenia spectrum disorders, or intellectual disability to control for comorbidities. Tier 3 further incorporated teacher-reported data from the Brief Problem Monitor (BPM) (Achenbach & Rescorla, 2001) to confirm symptom consistency across settings. Finally, Tier 4 applied clinical cutoffs from the parent-reported Child Behaviour Checklist (CBCL) attention or ADHD DSM-5 scale (Achenbach & Ruffle, 2000). The non-ADHD group included participants not labelled as ADHD by this tiered selection.

For the Lytle and colleagues’, we followed the study’s criteria, based on ADHD Rating Scale-IV (Pappas, 2006) and Parent-Kiddie Schedule for Affective Disorders and Schizophrenia for School-Age Children-Present and Lifetime Version (K-SADS-PL) (Hammer, Cooke, et al., 2015; Kaufman et al., 1997) (see Figure 1B).

### Cognitive functioning

Cognitive functioning was operationalised using age-appropriate, standardized composite measures available in each dataset. In the ABCD cohort, cognitive performance was indexed using the NIH Toolbox total cognition composite score (Bauer & Zelazo, 2013; Jernigan et al., 2024). In the external validation dataset from Lytle and colleagues, cognitive functioning was measured using the Full-Scale IQ score from the Wechsler Abbreviated Scale of Intelligence, Second Edition (WASI-II) (Lytle et al., 2020; Wechsler, 1999). Detailed descriptions of the component tasks contributing to each composite are provided in the Supplementary Methods.

To ensure numerical comparability across datasets, cognitive scores were z-score standardized within each cohort prior to model training and evaluation. To prevent data leakage, within each cross-validation fold, the mean and standard deviation of the cognitive score were calculated using the training participants only. These training-set parameters were then used to standardize both the training and corresponding test-set scores. This standardization places outcomes on a common scale and allows predictive performance to be assessed in terms of relative deviations within each population, rather than absolute score values tied to a specific instrument.

### Neuroimaging: ABCD

We analysed neuroimaging data from the Adolescent Brain Cognitive Development (ABCD-5.1) Study (Yang & Jernigan), combining pre-processed tabulated summary features provided by the ABCD consortium (Hagler et al., 2019) with additional custom processing of minimally pre-processed data from the ABCD-BIDS Community Collection (ABCC-2.0) (ABCD-BIDS Community). The ABCD tabulated data offer ready-to-use, quality-controlled summary measures and task contrasts parcellated with the Destrieux and FreeSurfer (Hagler et al., 2019). We complemented these with our own processing of ABCC data (using fMRIPrep 20.2.0 outputs and HCP minimal pre-processing pipelines) to derive higher-resolution region-to-region functional connectivity and task contrasts parcellated with the multimodal Glasser atlas (Glasser et al., 2016) (360 cortical regions) plus 19 FreeSurfer subcortical regions (Fischl et al., 2002) (total 379 regions). After applying ABCD-recommended quality control and exclusions (e.g., removing Site 22, correcting site ID errors, excluding participants with vision problems, and flagging poor-quality scans/tasks), the final sample for neuroimaging analyses included from 4000 to 11177 participants, depending on the modality and feature type. This resulted in 81 distinct sets of neuroimaging features (detailed in Supplementary Figure 1 and Supplementary Methods).

#### Functional MRI (fMRI) Task Contrasts: 56 sets

We derived task-based GLM contrasts from three fMRI tasks: Emotional N-back (working memory with emotional faces/places) (Barch et al., 2013), Monetary Incentive Delay (MID; reward/punishment processing) (Knutson et al., 2000), and Stop Signal Task (SST; inhibitory control) (Whelan et al., 2012). The ABCD tabulated data provided 26 pre-computed contrasts (9 N-back, 10 MID, 7 SST) parcellated with the Destrieux cortical (Destrieux et al., 2010) and FreeSurfer subcortical atlases (Hagler et al., 2019). Using the ABCC, we computed 30 contrasts (10 per task) via first-level General Linear Modelling (GLM) in Nilearn (see Supplementary Methods Supplementary Figure 1 for a complete list of contrasts). Pre-processing included nuisance regression (cosine drift regressors, 10 anatomical CompCor components, 6 motion parameters + derivatives), removal of non-steady-state volumes, and application of the SPM hemodynamic response function. Contrasts isolated specific cognitive processes (e.g., 2-back > 0-back for working memory load; large reward > neutral for reward magnitude; successful stop > go for inhibition). Effect sizes were averaged across runs (fixed-effects, variance-weighted) and parcellated into the 379-region atlas (Glasser cortical + FreeSurfer subcortical).

#### Functional MRI Connectivity (fMRI FC): 9 sets

Resting-state FC from ABCD tabulated data included three sets: (1) temporal variance (amplitude of low-frequency fluctuations) in 333 Gordon cortical parcels and 19 FreeSurfer subcortical regions; (2) subcortical-to-network FC (average correlation between each of 19 subcortical regions and 13 cortical networks from the Gordon (Gordon et al., 2016) parcellation); and (3) cortical network FC (mean correlations within and between those 13 networks; 91 features total). Using the ABCC, we computed region-to-region FC (Pearson correlation on residual time series after nuisance regression and, for task FC, regression of task events) at the 379-region resolution (Glasser cortical + FreeSurfer subcortical) during rest and each of the three tasks (four additional sets). We further derived multitask FC (concatenated residuals across all tasks) and general FC (concatenated residuals across rest + all tasks), following prior evidence that concatenated data improve reliability and brain-behaviour prediction (Cole et al., 2014, 2019; Elliott et al., 2019). Correlations were Fisher z-transformed and vectorized (see Supplementary Methods for more details).

#### Structural MRI (sMRI) and Diffusion Tensor Imaging (DTI): 16 sets

sMRI features (15 sets) included regional measures from FreeSurfer processing of T1-and T2-weighted images: cortical thickness, surface area, volume, sulcal depth, gray/white matter intensity, normalized intensities, and subcortical volumes (Destrieux cortical + FreeSurfer subcortical parcellations). Global “FreeSurfer summations” (one set) comprised five whole-brain metrics: estimated total intracranial volume, total cortical gray matter volume, total cortical white matter volume, total subcortical gray matter volume, and brain segmentation volume-to-intracranial volume ratio. Additional tabulated sets included T1/T2 summations (global intensities, differences between gray/white matter, cerebellar intensities, etc.) (Hagler et al., 2019). For DTI (one set), we used fractional anisotropy (FA) along 23 major white matter tracts (Atlas Track labelling), reflecting microstructural integrity and directional coherence of water diffusion (Hagler Jr et al., 2009). These structural features were selected to capture complementary aspects of brain morphology and white matter organization relevant to cognitive development.

### Neuroimaging: Lytle and colleagues’

Compared to the ABCD (Jernigan et al., 2024), Lytle and colleagues’ study (Lytle et al., 2020) provided much more limited neuroimaging modalities: two versions of the working-memory Nback task and sMRI (Hammer, Tennekoon, et al., 2015). See Supplementary Methodss for more details about how we processed these neuroimaging features.

Lytle and colleagues’ (2020) dataset provided more limited modalities than ABCD (Jernigan et al., 2024): two variants of the N-back working memory task (verbal and spatial, with reward/feedback manipulations) and sMRI. We processed these using the same first-level pipeline as for ABCC data (using fMRIPrep outputs, nuisance regression, GLM for contrasts, region-to-region FC on residuals at Glasser + FreeSurfer 379-region resolution). This yielded 13 sets of features (detailed in Supplementary Methods, Supplementary Figure 2 and Supplementary Methodss).

#### fMRI Task Contrasts: 6 sets

Unlike the ABCD, Lytle and colleagues used two versions of the working-memory Nback: verbal and spatial, and they also varied the reward size and feedback presence in each task (Hammer, Cooke, et al., 2015). We computed GLM contrasts from the two versions and averaged across the reward-size and feedback conditions using the same pre-processing and 379-region parcellation as ABCD. This left 3 contrasts for each version: 2back, 1back and 2-1back.

#### fMRI FC: 3 sets

Similar to the ABCD, we generated task FC by regressing out the HRF-convolved events. We then computed region-to-region (Glasser cortical + FreeSurfer) subcortical FC (Pearson correlation, Fisher z-transformed) separately for the verbal Nback, the spatial Nback, and the combined tasks as multitask FC. This dataset did not provide resting-state fMRI; consequently, the external-validation FC measures were task-derived rather than resting-state FC.

#### sMRI: 4 sets

We extracted cortical thickness, surface area, subcortical volume, and the same five FreeSurfer global summations described above.

### Cognitive prediction models

We predicted cognitive functioning scores from neuroimaging features using a two-layer modelling approach designed to evaluate predictive performance across individual sets of features and their combinations, while addressing high dimensionality, multicollinearity, and missing data. All models were trained and evaluated using a nested leave-one-site-out (LOSO) cross-validation scheme across the 21 ABCD sites to minimize site-specific biases and ensure generalizability (Cawley & Talbot, 2010). In the outer loop, each site was held out once as the test set, with the remaining 20 sites used for training. Both cognitive scores and features were z-scored using training-set statistics only (applied to the test set to prevent data leakage). In the inner loop, hyperparameters were tuned via five-fold cross-validation on the training data, optimizing negative mean squared error (NMSE).

#### Single models (81 models total)

For each of the 81 neuroimaging sets of features individually, we trained a separate predictive model as follows:

For functional connectivity sets (high-dimensional; ∼72,000 features per set), we first applied Partial Least Squares (PLS) regression as a supervised dimensionality reduction step to maximize covariance between features and the cognitive target (Krishnan et al., 2011). The number of PLS components was tuned via grid search (2–30 components maximum; all possible components considered for sets <30 features) using inner-fold NMSE.

The (reduced) features were then entered into Elastic Net linear regression (Zou & Hastie, 2005), which combines L1 (Lasso) and L2 (Ridge) regularization to promote sparsity and handle multicollinearity. Hyperparameters were tuned via grid search: regularization strength α (log-spaced from 10 ² to 10²) and L1-ratio (0 to 1 in 0.1 increments). All features in a given set were included in the model (no additional feature selection). The choice of Elastic Net was based on prior benchmarking study that showed Elastic Net to perform well in neuroimaging-based cognitive prediction (Pat et al., 2023). We also favoured these relatively parsimonious models over more highly parameterized alternatives because linear models have performed comparably to deep and kernel-based approaches for structural and functional brain prediction in large samples (Schulz et al., 2020), while prior cognitive-prediction work has found Elastic Net to perform similarly to or better than alternative algorithms (Tetereva et al., 2022, 2025). This choice also provides a comparatively computationally efficient modelling framework for the large number of feature sets evaluated here.

This resulted in 81 single-set models (one per feature set), each producing out-of-fold predictions on the held-out site.

#### Stacked models (15 + 5 configurations)

To integrate information across sets of features and capture potential non-linear interactions, we trained 20 stacked models using late fusion (predictions from single-set models as inputs, rather than raw feature concatenation). Each stacked configuration combined predictions from a related group of single-set models (e.g., "Stacked All" used predictions from all 81 single models; "stacked task contrasts" used predictions from the 56 task-based fMRI contrast sets; see Supplementary Methods Supplementary Figure 1 for full list). Of these 20 stacked models, five were assembled for external validation by combining predictions from single-set models corresponding closely to available sets of features in the external validation dataset.

The stacking regressor was a Random Forest (Breiman, 2001) with 1,000 trees. Hyperparameters were tuned via inner-fold grid search: maximum tree depth (None or 3–10), and number of features considered at each split (sqrt or log2 of total input features). Random Forest was chosen for the stacking layer because it naturally handles mixed-scale inputs, non-linear relationships, and interactions between modalities without requiring further normalization (Tetereva et al., 2022).

Missing predictions in the stacking layer (due to incomplete neuroimaging data for some participants or tasks) were addressed using opportunistic stacking (Engemann et al., 2020; Pat et al., 2022). For each single-model prediction, we created two input features: one with missing values imputed as +1000 and one as –1000. This retained participants with partial data (as long as at least one feature set was available) while explicitly signalling missingness to the Random Forest. Using the ABCD dataset, Pat et al. (2022) demonstrated that opportunistic stacking achieved predictive performance comparable to listwise deletion while substantially increasing the number of participants that could be included in the analysis.

#### Feature importance

Feature importance, also known as algorithmic importance, was quantified using model-specific measures appropriate to each predictive architecture, reflecting how each model utilised information to generate predictions. For white-box models, such as Elastic Net and PLS, their coefficients are readily interpretable. For Elastic Net, feature importance was defined directly by the fitted regression coefficients, reflecting the direction and magnitude of each feature’s association with the predictions. For PLS that was applied to FC, importance was derived from the fitted PLS regression model. Specifically, we computed feature weights by combining the PLS X-component loadings with the predictive contribution of each component, quantified as the out-of-sample variance explained (R²) by that component. The final FC feature importance score was obtained as the dot product between the matrix of PLS feature weights and the vector of component-wise explained variance, yielding a single variance-weighted importance value per pair. For stacked ensemble models based on Random Forests, feature importance was estimated using the algorithm-agnostic Shapley Additive exPlanations (SHAP) framework to assess how model predictions change when a feature is added across all possible combinations of the remaining features. SHAP values were computed within each cross-validation fold, aggregated across subjects by taking the mean absolute SHAP value per feature, and then averaged across folds to obtain a robust global ranking of feature contributions (Bifarin, 2023; Lundberg & Lee, 2017). Because these are multivariate measures, their values reflect algorithmic importance conditional on the other predictors and their correlation structure, rather than independent biological effects or causal neurobiological importance (Haufe et al., 2014).

### Application to external validation sample

Due to the differences in feature availability between the ABCD and Lytle and colleagues’, we were restricted to the sets of features that had corresponding or approximately comparable representations in both datasets. Specifically, we used single and stacked models trained on a subset of Nback contrasts (0back, 2back and 2-0back), which manipulated working memory load—collectively referred to as working-memory (WM) Load contrasts for the stacked model trained on these three—along with Nback FC, multitask FC, general FC, and sMRI data from the ABCD. These models were then applied to single and stacked sets of features, including verbal and spatial Nback contrasts (1-back, 2-back, and 2-1-back), as well as verbal and spatial Nback FC, multitask (verbal + spatial) FC, and sMRI data, in Lytle and colleagues’ study. Supplementary Table 1 shows which models trained on the ABCD are applied to which sets of features in Lytle and colleagues’ dataset). Because resting-state fMRI was unavailable in external validation, the ABCD general FC model was mapped to the closest available multitask FC representation; task-event regression removes the modelled task-evoked component while retaining connectivity with substantial similarity to intrinsic network organisation (Cole et al., 2014; Elliott et al., 2019). Thus, external validation represents a targeted test of models for which comparable features were available, not a validation of all 81 ABCD feature sets.

### Potential confounding and sensitivity analysis

Several steps were taken to address potential confounding. Leave-one-site-out cross-validation was used to account for site-related variation, while image quality and head motion were addressed during neuroimaging preprocessing and quality control. ABCD-derived features followed ABCD-recommended quality-control flags (including automated and manual quality-control criteria as well as task-performance checks), whereas ABCC-derived fMRI analyses included nuisance regression using anatomical CompCor components and motion parameters. The ADHD tiering procedure also excluded several major comorbid conditions at Tier 2.

Nevertheless, demographic and medication-related factors may still influence both cognitive performance and model predictions. To evaluate their potential impact, we conducted a post-hoc sensitivity analysis in which both observed and predicted cognitive scores were residualised for sex, parental education, household income, and medication exposure. Medication exposure was classified into 14 main groups based on the first-level categories of the Anatomical Therapeutic Chemical (ATC) system. Prediction performance was then recalculated using the residualised scores. This analysis was designed to assess the extent to which demographic, socioeconomic, and medication-related factors contributed to the predictive performance found in each ADHD group. Further details are provided in the Supplementary Methods.

### Evaluation

Correlation (r), coefficient of determination (R²), mean absolute error (MAE), and root mean squared error (RMSE) were reported as measures of predictive performance across all folds for internal-validation-set and for whole external-validation-set, as well as separately for ADHD and non-ADHD subgroups.

To evaluate whether the model with the highest nominal predictive accuracy reliably outperformed alternative models, we conducted pairwise comparisons between the highest nominal model (reference model) and every other model. Accounting for differences in sample size, predictions were aligned at the subject level by merging test-set predictions across the two models using participant IDs. We then performed bootstrap resampling with replacement across subjects (n = 2000 iterations). In each bootstrap iteration, we computed the difference in predictive correlation between models such that positive values indicate improved predictive accuracy for the reference model. Bootstrap 95% confidence intervals were obtained using the percentile method (2.5th and 97.5th percentiles). A performance difference was considered statistically significant when the confidence interval did not include zero. Model comparisons were performed separately for internal validation and external validation.

Group differences in performance metrics (MAE, RMSE, R², and Pearson r) between ADHD and non-ADHD participants were evaluated using percentile bootstrapping (2,000 iterations). Within each iteration, participants were resampled with replacement independently within each group to a fixed balanced size (1,000 per group in ABCD; 200 per group in the Lytle dataset). The metric was computed separately for each resampled group, and the difference (ADHD minus non-ADHD) was stored. The 95% confidence interval for each difference was derived from the 2.5th and 97.5th percentiles of the bootstrap distribution of differences; differences were considered statistically significant if the CI excluded zero.

Confidence intervals for Pearson *r* (within groups or overall) were computed using percentile bootstrapping (1,000 iterations). In each iteration, paired observed and predicted values were resampled with replacement (preserving the original sample size), the correlation was recalculated, and the 95% CI was taken as the 2.5th and 97.5th percentiles of the resulting distribution.

Differences in the distributions of observed and predicted cognitive scores between non-ADHD participants and ADHD subgroups (Tiers 1–4 in ABCD; single ADHD group in Lytle et al.) were evaluated by comparing group means. Welch’s independent-samples t-test (unequal variances assumed) was first performed for each score type. Effect sizes were quantified using Hedges’ *g*, computed as Cohen’s *d* based on the pooled standard deviation (s_pooled) and corrected for small-sample bias. Cohen’s *d* was defined as:

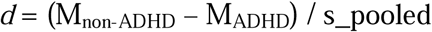

and Hedges’ g was obtained by applying the standard correction factor:

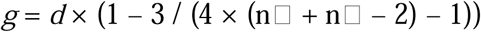

Degrees of freedom were estimated using the Welch–Satterthwaite approximation. Uncertainty in effect size estimates was quantified by computing 95% confidence intervals for Hedges’ *g* using a normal-approximation standard error.

See Figure 1 C&D and Supplementary Methods for more details about the prediction models and evaluation procedure. All utilized code is available in the public domain at https://github.com/HAM-lab-Otago-University/ADHD-MultiModal.

## RESULTS

### Group differences in cognitive functioning scores: observed vs. model-predicted

To assess whether neuroimaging-based predictions reproduced group differences in observed cognitive functioning, we quantified the separation between non-ADHD and each ADHD tier in both (i) observed cognitive functioning scores and (ii) model-predicted cognitive functioning scores.

#### Internal Validation

Figure 2A shows whether model predictions reproduce observed ADHD/non-ADHD group differences. Observed cognitive functioning scores were statistically significantly higher for non-ADHD compared to each ADHD tier. Predicted scores from the best-performing Stacked All model replicated this pattern, with predicted scores higher for non-ADHD than each ADHD tier, and with effect sizes comparable in magnitude to those computed from the observed scores (Figure 2A). As shown in Figure 2B (Hedges’ *g*), most high-ranked models—particularly stacked models—showed predicted-score group differences that mirrored the observed pattern, with effect sizes closely tracking those derived from observed scores. In contrast, lower-ranked models more frequently yielded attenuated or inconsistent group separation.

**Figure 2.**
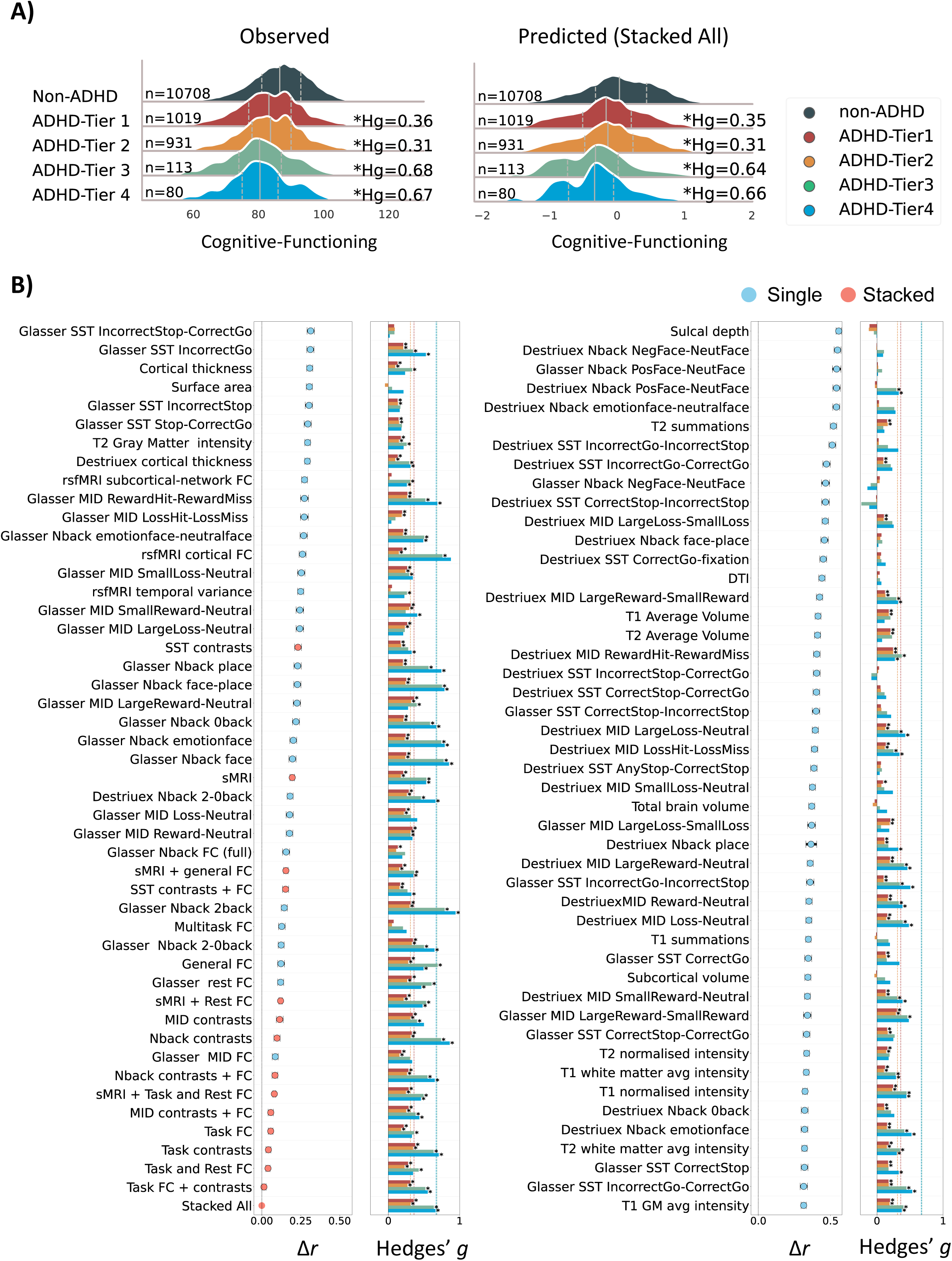
(A) Distributions of observed and predicted cognitive functioning scores in the internal validation set. Predicted scores are generated by the Stacked All model, which achieved the highest overall predictive performance (Pearson’s r). **(B) Left panels: Difference in Pearson’s correlation (**Δ**r) between Stacked All and each model.** Larger values indicate worse performance compared to the Stacked All model. Bars indicate bootstrap 95% confidence intervals. **Right panels: Effect sizes (Hedges’ g) for differences in predicted cognitive scores between the non-ADHD group and each ADHD tier, shown separately for each model.** Vertical lines indicate the corresponding effect size computed from observed cognitive scores. Asterisks denote statistically significant differences between the non-ADHD group and each ADHD tier.

#### External Validation

As shown in Figure 3A, observed cognitive functioning scores were also statistically significantly higher for non-ADHD than ADHD. Predicted scores derived from the highest nominal external validation model (stacked spatial Nback working-memory load contrasts) showed a corresponding non-ADHD > ADHD shift, although effect sizes were numerically smaller than those obtained from observed scores (Figure 3A).

**Figure 3.**
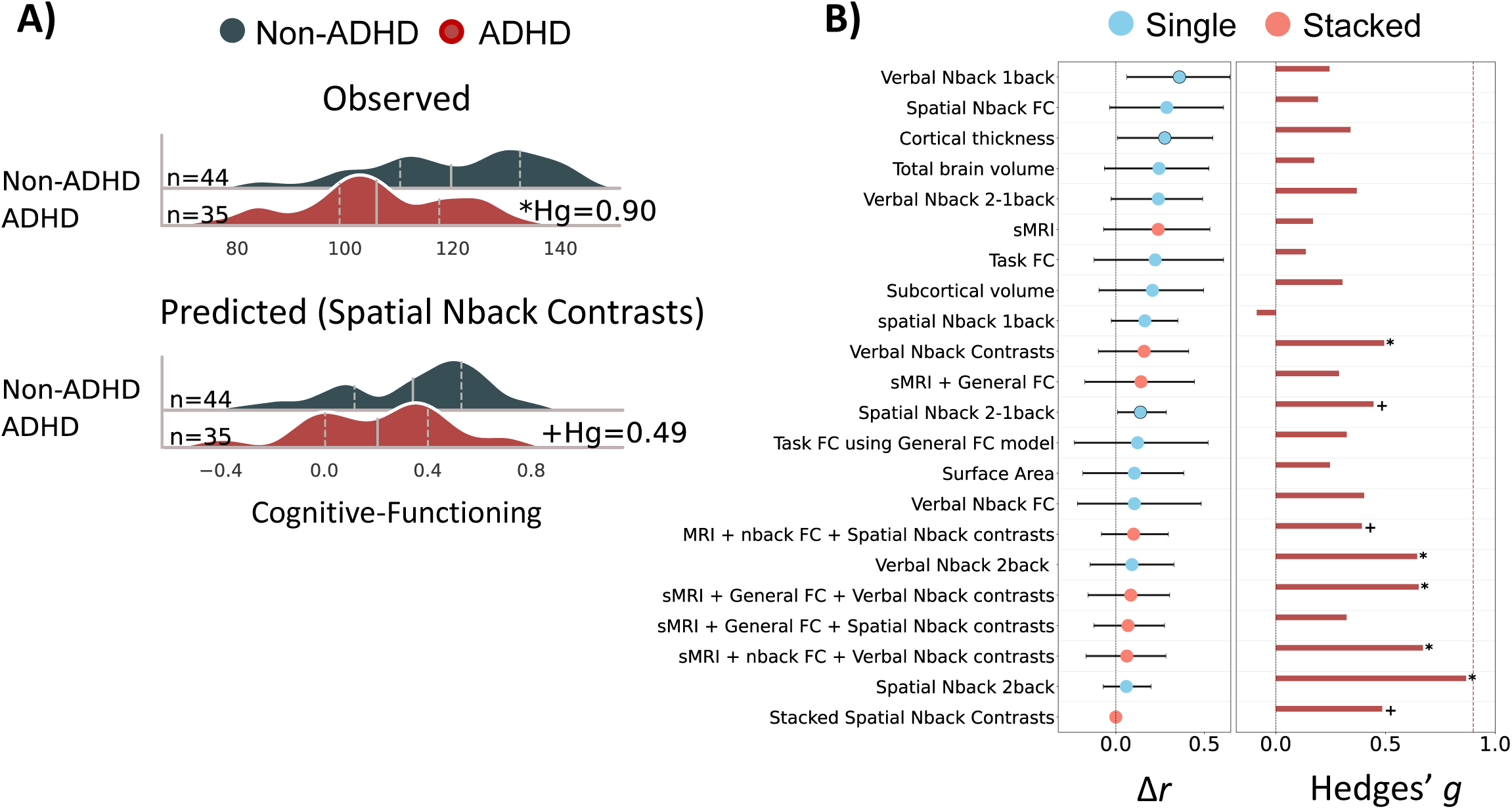
(A) Distributions of observed and predicted cognitive functioning scores in the external validation set. Predicted scores are generated by the stacked model, which achieved the highest nominal overall predictive performance (Pearson’s r). **(B) Left panel: Difference in Pearson’s correlation (**Δ**r) between the top stacked and each model.** Larger values indicate worse performance compared to the top Stacked model. Bars indicate bootstrap 95% confidence intervals. **Right panel: Effect sizes (Hedges’ g) for difference in predicted cognitive scores between the non-ADHD group and ADHD group, shown separately for each model.** The vertical line indicates the corresponding effect size computed from observed cognitive scores. Asterisks/plus denote statistically/partially significant differences between the non-ADHD and ADHD group.

Across models, effect size summaries indicated that models with higher predictive performance (i.e., numerically larger *r* values) reproduced the observed group difference, but with attenuated magnitudes and more variable statistical significance. This was reflected in both the ridge density plot (Figure 3A) and the Hedges’ *g* bar plots across models (Figure 3B, Hedges’ *g*). Lower-ranked models showed weaker and less consistent separation.

### Predictive accuracy and comparison of stacked vs. single models

#### Internal validation

Across all participants (regardless of diagnostic group), the stacked model integrating predictions from all 81 neuroimaging sets of features (Stacked All) achieved the highest out-of-sample predictive accuracy, with an overall correlation of r = .57 (95% CI = [.56–.59]). We compared the Stacked All model against every other stacked and single model using aligned-subject bootstrap resampling. These analyses indicated that Stacked All significantly outperformed all other models in internal validation (Figure 2B, Δr).

Among the single models, task FC, rest FC, and the Nback 2–0back contrast showed the nominally highest predictive accuracy (Figure 2B, Δr).

Sensitivity to demographic, socioeconomic, and medication-related confounding was assessed by residualizing both observed and predicted cognitive scores for parental education, household income, sex, and medication exposure. Predictive performance for the Stacked All model after residualization is reported in Supplementary Table 2. Overall, residualizing parental education and household income modestly attenuated model performance, whereas residualizing sex and medication exposure had little effect. Importantly, any attenuation was consistent across ADHD status. For example, in the Stacked All model, the predictive correlation (r) in the non-ADHD group decreased from .57 to .50 after adjusting for parental education and to .47 after adjusting for household income, but remained essentially unchanged after adjusting for sex (r = .57) or medication exposure (r = .57). Similar patterns were observed across ADHD Tiers 1-4.

#### External validation

In external validation, the stacked Nback working-memory load contrast model applied to spatial Nback contrasts (2back, 0back, and 2–0back) achieved the highest nominal predictive accuracy, with r = .44 (95% CI = [.19–.52]). This performance closely matched its internal validation performance (r = .45; 95% CI = [.46–.50]). The second-highest nominal model was trained on all applicable sets of features (sMRI, general FC, and Nback WM load contrasts) and applied to sMRI, multitask FC, and spatial Nback contrasts, achieving r = .37 (95% CI = [.03–.41]). Aligned-subject bootstrap comparisons showed that, unlike internal validation, the highest nominal external validation model did not consistently outperform the majority of other models, although it remained numerically higher than competing models (Figure 3B, Δr).

The Supplementary Tables 3-6 include the r, R², MAE, and RMSE values for ADHD and non-ADHD subgroups in internal and external validation.

### Predictive Performance Across Groups

#### Internal Validation

Bootstrapping analyses did not reveal any statistically significant differences in predictive performance between non-ADHD and any ADHD tiers for the Stacked All model. This comparable performance across non-ADHD and ADHD of any tiers held for 10 of 15 stacked models (Figure 4A) and 27 of 81 single models (Figure 5A). Notably, unlike FC stacked models (e.g., All FCs, task FCs, rest FCs), task contrasts models exhibited reduced predictability in stricter ADHD tiers (e.g., Tiers 3 or 4) compared to non-ADHD (e.g., Nback Contrasts: Δr_ADHD-Tier4-vs-non-ADHD_= -.11; 95% CI = [-0.18-0.04]). In the bootstrap correlation plots, stacked models more frequently retained correlations clearly above zero in stricter ADHD tiers (Tiers 3–4), whereas many single models showed confidence intervals crossing *r* = 0, (Figure 4A; Figure 5A). Figure 6A illustrates the correlation between observed and predicted cognitive functioning scores for the nominally best stacked and single models across task contrast, FC, and sMRI sets of features.

**Figure 4.**
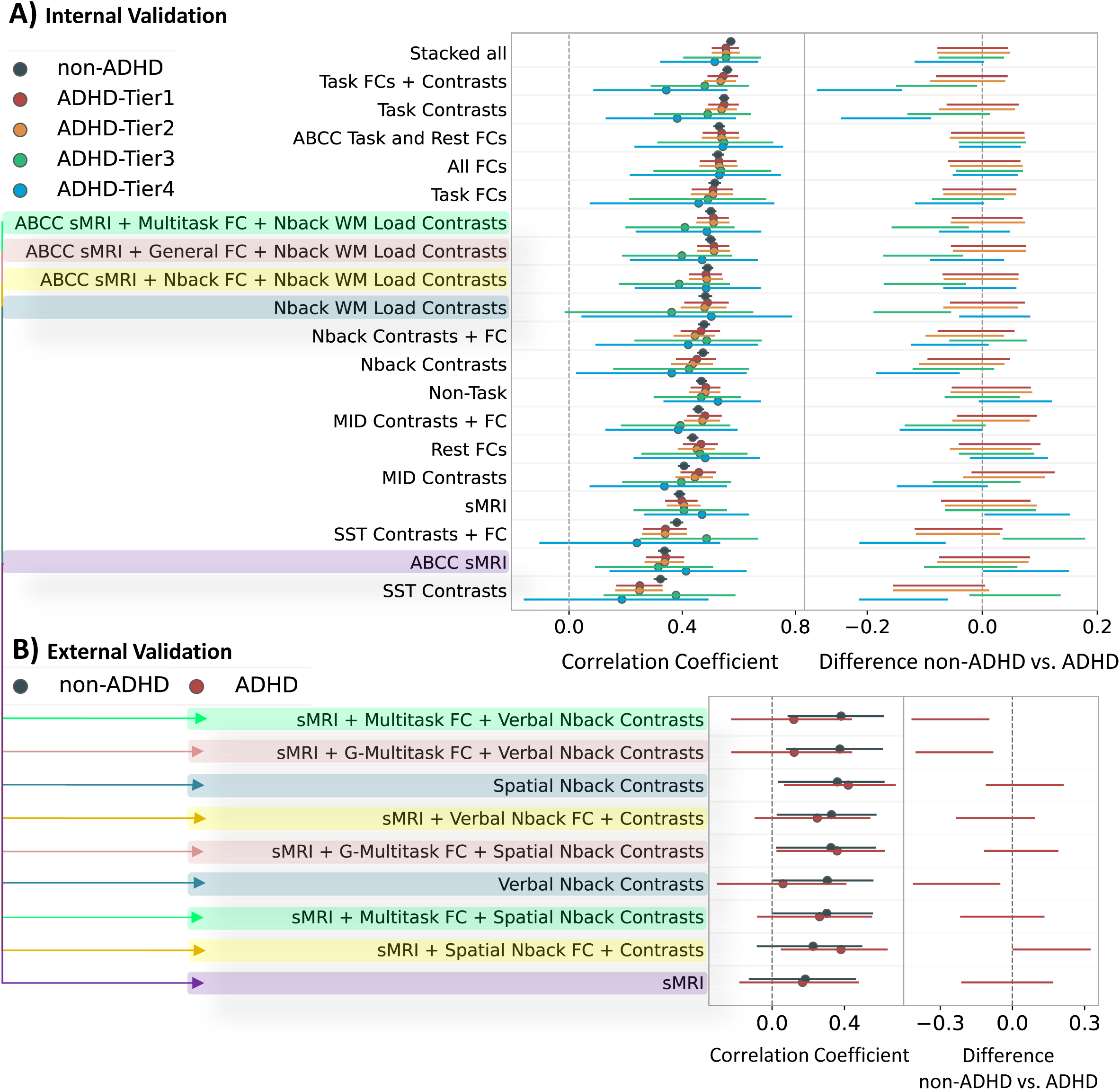
Predictive performance of stacked models across groups in (A) internal and (B) external validation datasets, ranked by performance in non-ADHD groups. The left panel shows Pearson’s correlation between predicted and observed values for cognitive functioning scores averaged across sites. The bars show bootstrapped 95% CI. The right panel shows the bootstrapped confidence interval for differences in model performance between non-ADHD and each ADHD group. Intervals below/above zero indicate statistically significantly worse/better performance in ADHD groups at 95% CI, respectively. The highlighted colours indicate which stacked models from the internal-validation set were applied to which stacked sets of features in the external-validation set. Five stacked models from the internal validation set were applied to the external validation set, including ABCC sMRI, WM load contrasts from the Nback task and their combination with Nback, multitask or general FCs. The four stacked models that included task-related features were applied separately to verbal and spatial Nback tasks in the external-validation set, yielding in total nine stacked models for the external-validation set. The G-prefix for multitask FC refers to the models trained on general FC in the ABCD that were then applied to multitask FC in the external-validation set. Although 15 stacked configurations were defined for internal validation, only nine stacked model applications are shown in the external-validation panel because only five configurations had compatible feature sets and four of these were evaluated separately on the verbal and spatial N-back tasks.

**Figure 5.**
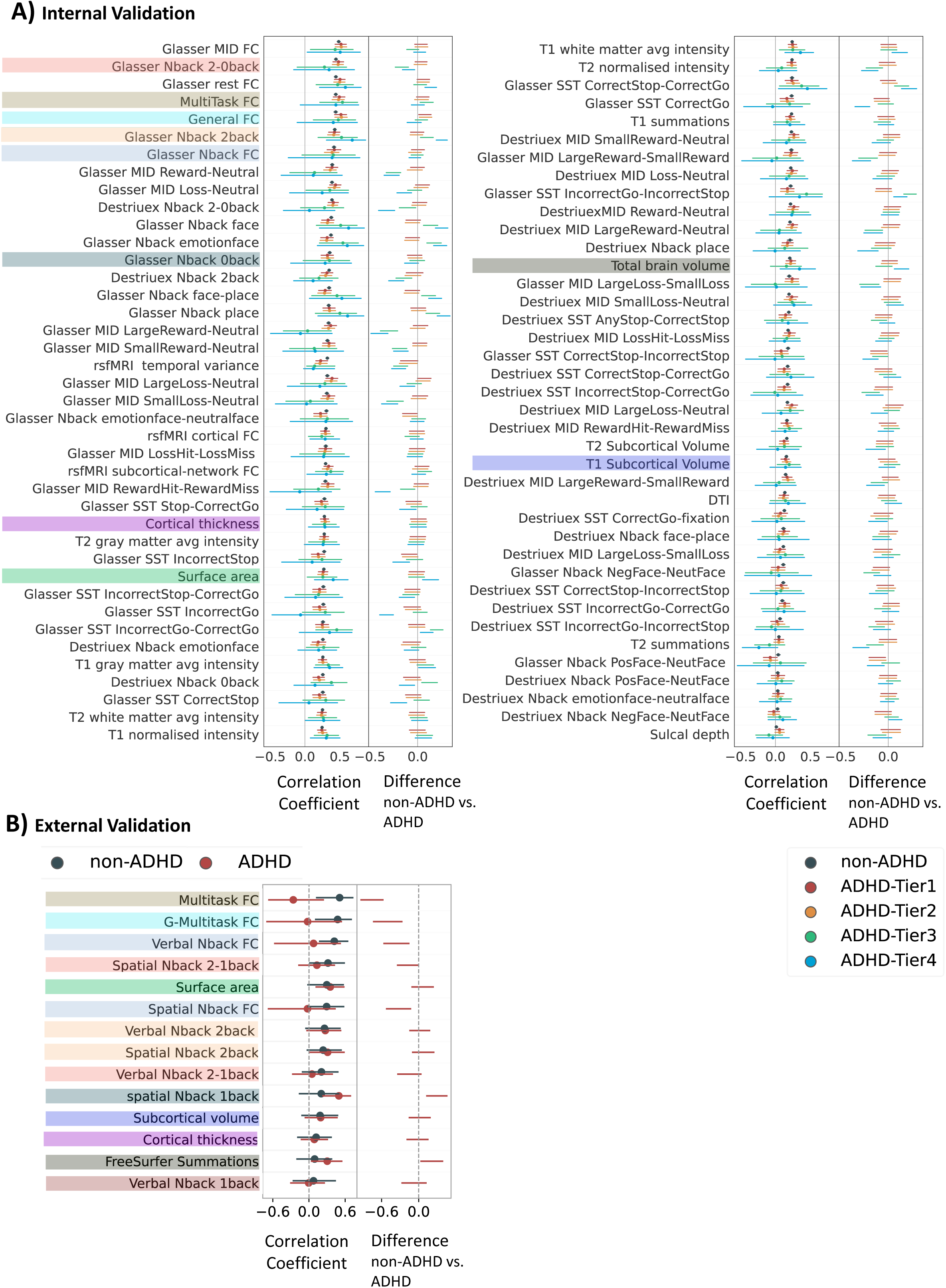
Predictive performance of single models across groups in (A) internal and (B) external validation, ranked by performance in non-ADHD groups. The left panel shows Pearson’s correlation between observed and predicted values for cognitive-functioning total scores. The bars show bootstrapped 95% CI. The right panel shows the bootstrapped confidence interval for the differences in model performance between non-ADHD and ADHD tiers/statuses. intervals below/above zero show significantly worse/better performance in ADHD tiers/statuses, respectively. The highlighted colours indicate which models from the internal-validation set were applied to which sets of features in the external-validation set.

**Figure 6.**
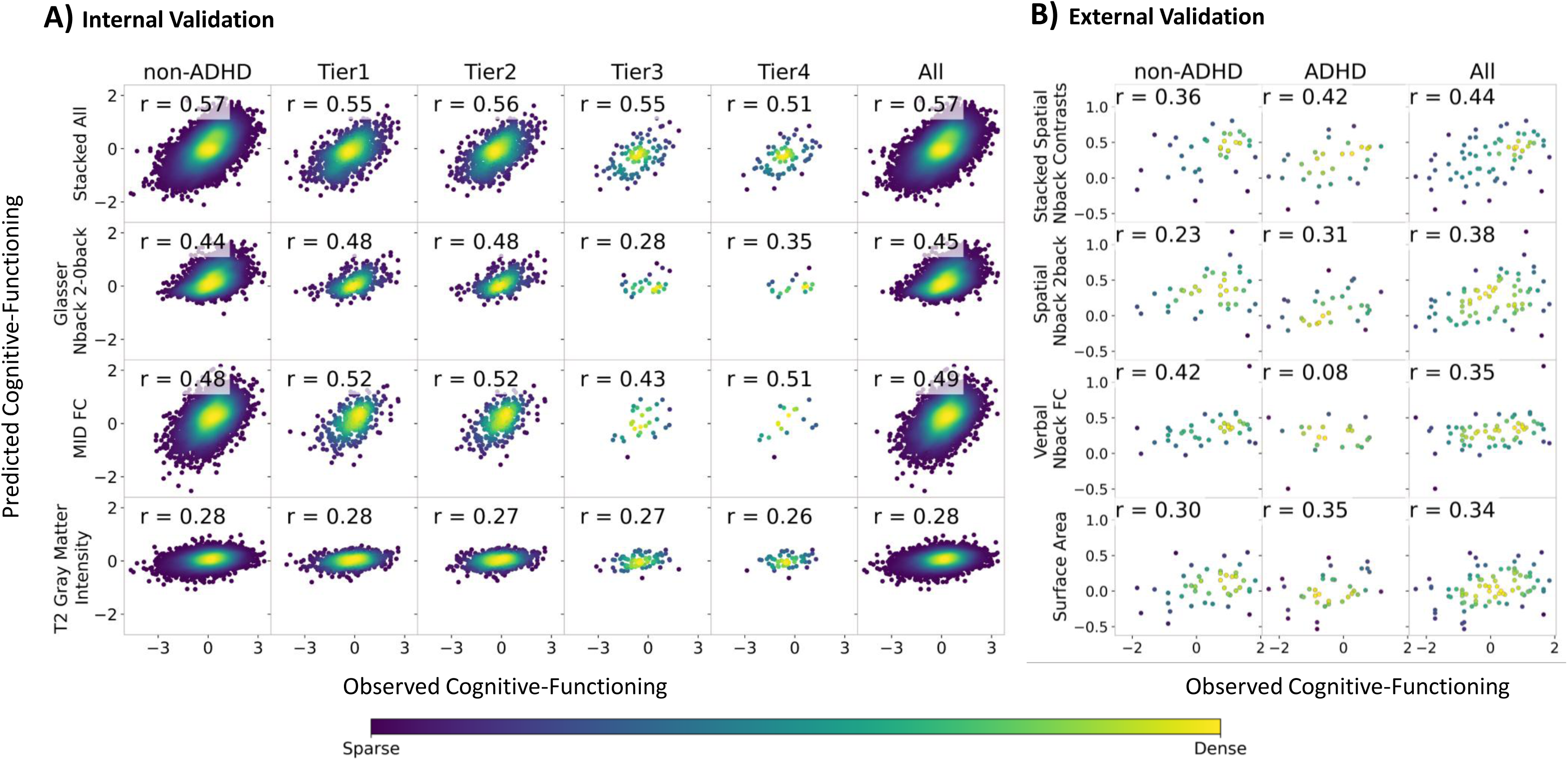
The scatterplots of the correlation between observed and predicted cognitive functioning scores for the nominally best stacked model and the best contrast-based, FC-based, and sMRI-DTI-based single models, in (A) internal and (B) external validation sets. Glasser Nback 2-0back indicates fMRI task contrast from the Emotional Nback task, which was parcellated using the Glasser atlas (Glasser et al., 2016). Glasser MID FC indicates task FC during the Monetary Incentive Delay (MID) task, which was parcellated using the Glasser atlas (Glasser et al., 2016).

#### External Validation

Neither of the two nominally highest stacked models showed statistically significant differences in predictive performance between ADHD and non-ADHD groups (see Figure 4B). Among single sets of features, the spatial and verbal 2back showed highest correlation values (*r* > .35 for both), with no significant difference revealed across groups (Figure 5B). Figure 6B displays the correlation between observed and predicted cognitive functioning scores for the nominally best stacked and single models in each measurement set.

### Feature Importance

Feature importance analysis (Figure 7) highlights key contributors to predicting cognitive functioning scores in the nominally best-performing models. In the stacked-all model that included every neuroimaging set of features, the Nback 2–0back contrast and task FC showed the largest average marginal contributions to the model’s predictions, as reflected by absolute SHAP values (Nback: M = .30, SD = .002; task FC: M = .14, SD = .05; Figure 7A). Among single models, task contrasts parcellated with the Glasser atlas achieved higher predictive performance (mean correlation between observed and predicted values) than those based on the Destrieux atlas (Supplementary Tables Supplementary Table 3). However, Destrieux-based contrasts ranked higher in the stacked model’s SHAP-based importance ranking, indicating that they provided additional predictive information when combined with other modalities.

**Figure 7.**
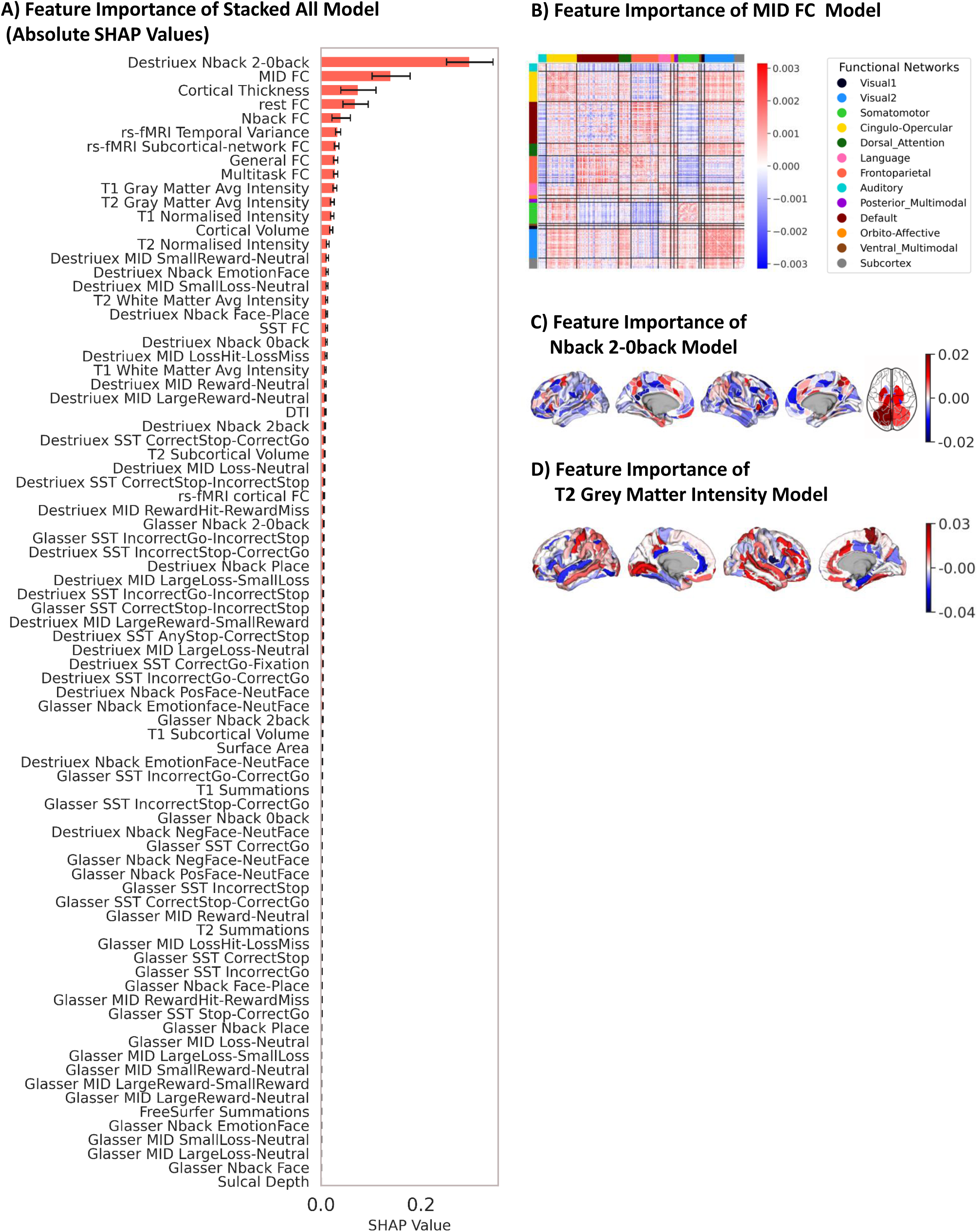
Feature Importance of stacked and single models. **(A)** Feature importance of the Stacked All model that included all sets of neuroimaging features. This feature importance is based on absolute SHAP values, averaged across test sites in the ABCD. A higher absolute SHAP value indicates a higher contribution to the prediction. Error bars reflect standard deviations across sites. **(B)** Feature importance of the nominally best model among FC models: the MID FC. - Feature importance of FC is based on the weight of each feature in the PLS components and the amount of variance explained by each component average across sites, visualized using the Cole-Anticevic Brain-wide Network Partition (Cole et al., 2024; Ji et al., 2019). **(C)** Feature importance of the nominally best-performing model among task contrasts: the Nback 2-0back model. Feature importance of task contrast is based on Elastic Net coefficients, averaged across sites. **(D)** Feature importance for the nominally best model among sMRI and DTI: T2 Grey Matter Intensity. Feature importance of sMRI and DTI is based on Elastic Net coefficients, averaged across sites.

For the top-ranked FC single model with (MID FC), the highest positive predictive weights were observed for connections involving the Dorsal Attention and Posterior Multimodal networks (Supplementary Tables Supplementary Table 7). For the top-ranked single contrast model (Nback 2–0back), Elastic Net coefficients placed the strongest positive predictive weights on ventral occipital and anterior cingulate cortex (ACC) regions (Supplementary Tables Supplementary Table 8). For the top-ranked structural model (T2 grey matter intensity), the largest positive predictive weights were observed for the Paracentral Gyrus and Sulcus and the Cingulate Marginal Sulcus (Supplementary Tables Supplementary Table 9). These coefficient- and weight-based patterns should be interpreted as reflecting how the fitted multivariate models combined features to optimize prediction, rather than as direct evidence that the highlighted regions exhibit the strongest association with cognitive functioning.

## DISCUSSION

This study provides the first direct test of whether multimodal cognitive prediction models, trained on a large-scale, population-based cohort, can generalize to clinical population—in this case, children with ADHD—without any dataset-specific tuning. Prior work examining model generalizability to psychiatric conditions has largely relied on single-modality neuroimaging (e.g., structural MRI or resting-state fMRI alone) or has introduced intermediate adaptation steps that limit conclusions about true out-of-sample performance (Chopra et al., 2024; Samea et al., 2019, 2019; Seidman et al., 2005). Equally important, our work provides a comprehensive exploratory benchmark of model generalizability across 81 MRI-derived feature types, spanning structural, diffusion, task-based activation, and functional connectivity measures. This systematic evaluation offers valuable insight into which modalities—and combinations thereof—are most informative for cognitive prediction in ADHD and non-ADHD groups.

### Predicted scores reflect observed group differences across models

Across both internal and external datasets, cognitive functioning differed between the non-ADHD group and ADHD tiers, consistent with prior evidence of cognitive variation in ADHD populations (Duff & Sulla, 2015; Frazier et al., 2004; Kofler et al., 2019; Pievsky & McGrath, 2018). Critically, model predictions reproduced these group-level patterns: across most high-performing models, predicted cognitive scores preserved the direction of observed group differences, and effect sizes derived from predicted values closely tracked those derived from observed scores. This indicates that model outputs preserved group-level distributional differences in cognition. The magnitude of these effects should nevertheless be interpreted as group-level evidence. The effect sizes are broadly comparable to the mean neurocognitive effect size reported across ADHD meta-analyses (approximately g/d = .45; (Pievsky & McGrath, 2018), Importantly, this pattern was not restricted to a single best model. Rather, it emerged across a set of top-ranked models, supporting the conclusion that multiple neuroimaging feature types contain reproducible signal related to cognitive functioning.

### Generalization across ADHD tiers

Despite this distributional difference in cognitive scores, the majority of stacked models showed comparable performance between ADHD and non-ADHD groups. Within the present study, this pattern provides evidence for a brain–cognition association that is shared across ADHD and non-ADHD groups. This generalizability suggests that integrating multimodal neuroimaging features—such as task-based fMRI contrasts, functional connectivity, and structural MRI—may represents variability in cognitive processes like attention, working memory, and executive function, which are relevant to ADHD but not exclusive to its deficits (Abramovitch et al., 2021; East-Richard et al., 2020; Kofler et al., 2019; McTeague et al., 2016; Morris & Cuthbert, 2012). This aligns with prior studies using single-modality neuroimaging to predict cognitive performance in ADHD (Cortese et al., 2021; Rosenberg et al., 2016; Samea et al., 2019).

### Benefit of Multimodal Integration

In addition to comparable performance across non-ADHD and ADHD, stacking enhanced prediction accuracy over single sets of features, and tended to yield more stable performance across diagnostic tiers than single models. In contrast, several single models showed weaker or inconsistent subgroup prediction, including more unstable generalization across tiers. This suggests that multimodal integration may reduce sensitivity to modality-specific noise or subgroup-specific variability, resulting in more robust performance when models are applied across heterogeneous clinical strata. Task-based fMRI contrasts reflect activation patterns during cognitive demands, while FC captures intrinsic network dynamics, and structural features provide insights into brain morphology that may influence cognitive capacity (Litwińczuk et al., 2023; Mather et al., 2013; Zimmermann et al., 2018). By combining complementary neural signals, stacked models captured a richer representation of cognition-related processes which are critical for cognitive functioning across healthy populations (Barch et al., 2013; Elliott et al., 2019; Marek et al., 2022; Rasero et al., 2021; Rosenberg et al., 2018; Tetereva et al., 2022, 2025) and those with ADHD (Cordova et al., 2022; Frazier et al., 2004; Pievsky & McGrath, 2018). This multimodal synergy enhances the models’ ability to generalize to ADHD populations, where cognitive functioning varies widely but is not exclusively tied to disorder-specific alterations in particular brain regions, networks, or structures (Cortese et al., 2021; Luders et al., 2009; Mooney et al., 2024; Norman et al., 2016; Seidman et al., 2005).

### Residualization Sensitivity Analyses

The residualization sensitivity analyses indicated that the Stacked All model was robust to adjustment for sex and medication exposure. However, adjusting for parental education and household income numerically reduced predictive performance, with the the Stacked All model’s correlation decreasing from r = .57 to r = .47-.51. Critically, the magnitude of attenuation was comparable across ADHD and non-ADHD groups, indicating that these covariates are unlikely to explain differences in predictive performance across ADHD status.

The observed influence of sociodemographic factors is consistent with prior research demonstrating associations between socioeconomic status and cognitive functioning in children and adolescents (Pat et al., 2022; Pievsky & McGrath, 2018; Rakesh et al., 2024; Vrantsidis et al., 2020). Notably, Pat and colleagues (2022) applied a similar stacking approach in the ABCD study and found that the stacked brain biomarker mediated 18.65% of the association between sociodemographic factors and cognitive performance. Accordingly, we do not view sociodemographic factors as confounders per se, but rather as factors that are intrinsically linked to both brain development and cognitive functioning.

### External validation: a stringent test across cohorts, tasks, and cognitive instruments

External validation provided a stringent test of transferability because it involved differences in samples, tasks, imaging features, and cognitive measures. Stacked Nback models showed similar correlation values across datasets, providing preliminary evidence that some cognition-related neural information can transfer across contexts (Chopra et al., 2024). Notably, we applied trained models directly to the external dataset without adaptation—a stricter test of generalizability relevant to clinical settings where model tuning may be impractical. Models trained on ABCD working memory Nback contrasts performed better when applied to the spatial Nback task in the external dataset than to the verbal Nback task, likely reflecting closer alignment in task demands. This task-specific pattern suggests that although models capture cognition-related neural activity, similarity in cognitive paradigms can meaningfully influence predictive accuracy. Importantly, the external validation should be regarded as preliminary rather than as evidence of robust generalization: the external validation does not assume equivalence between NIH Toolbox and WASI-II scores at the level of individual tasks or raw metrics. Instead, it rests on the assumption that both instruments provide valid, though not identical, operationalizations of a shared latent construct—general cognitive functioning—encompassing overlapping domains such as verbal ability, working memory, executive function, processing speed, and reasoning. Under this framing, external validation evaluates whether neuroimaging models trained to predict NIH Toolbox–based cognition in ABCD generalize to a different cohort in which the same underlying construct is assessed using an alternative psychometric instrument. Consequently, model performance in external validation reflects not only generalisation across samples and scanners, but also robustness to differences in cognitive measurement. This cross-instrument mismatch likely contributes to the reduced predictive accuracy observed in external validation and means that performance reflects transfer at the level of the broader construct of cognitive functioning rather than prediction of identical psychometric scores.

### Feature importance

We computed feature-importance (algorithm importance) estimates for each algorithm to characterize how the multivariate models leveraged information from different brain indices. Across both the Stacked All model (using SHAP-based feature importance) and the correlation-based rankings of the single-modality models, task-based N-back contrasts and functional connectivity measures emerged as consistently prominent predictors. This finding aligns with previous research highlighting the predictive utility of these measures for cognitive functioning (Chopra et al., 2024; Finn et al., 2015; Rosenberg et al., 2016; Sripada et al., 2020; Tetereva et al., 2025; Wang et al., 2025). Specifically, SHAP analyses (Bifarin, 2023; Lundberg & Lee, 2017) indicated that predictions from the Stacked All model were most strongly influenced by the inclusion of task-based N-back contrasts and functional connectivity measures across all possible combinations of the remaining modalities, suggesting that these features contributed substantially to predictive performance.

The prominence of N-back contrasts is consistent with their role in indexing working memory load and cognitive control, as they capture neural activation patterns within prefrontal and parietal regions that support attention and executive functioning (Owen et al., 2005). Similarly, functional connectivity, particularly task-based functional connectivity, reflects network-level interactions that underpin cognitive flexibility, sustained attention, and goal-directed behavior (Elliott et al., 2019; Stevens & Spreng, 2014; Zimmermann et al., 2018). Within these modalities, the identified importance of Dorsal Attention-Posterior Multimodal and Ventral Multimodal connectivity patterns (as reflected by PLS weights and loadings), together with anterior cingulate and ventral occipital activation during the N-back task (as reflected by Elastic Net coefficients), is broadly consistent with the established roles of these regions and networks in attentional control, sensory integration, semantic processing, and cognitive and affective regulation (Bush et al., 2000; Castellanos & Proal, 2012; Dosenbach et al., 2008; Osher et al., 2019; Power et al., 2011; Rosenberg et al., 2016; Shulman et al., 2009).

Importantly, these feature-importance findings should be interpreted as reflecting the contribution of features to model prediction rather than providing direct evidence of neurobiological importance or causal mechanisms underlying cognitive functioning (Haufe et al., 2014). For instance, Elastic Net feature importance depends not only on predictive relevance but also on how the algorithm allocates information across correlated features. When the solution is closer to Lasso, one predictor from a set of highly correlated brain indices may be selected while the others are shrunk toward zero. In contrast, when the solution is closer to Ridge, multiple correlated predictors are more likely to be retained, albeit with smaller coefficients. Consequently, estimated feature importance reflects both the predictive value of a feature and the algorithm-specific manner in which information is distributed across the multivariate feature space.

### Practical implications

Our benchmarking highlights a set of potentially high-yield sets of features—particularly working memory–related task contrasts and functional connectivity metrics—that consistently predict cognitive functioning across groups and datasets. These results may inform more efficient neuroimaging protocols by prioritizing acquisition of features most likely to yield robust brain–cognition prediction, potentially reducing scan time and cost without sacrificing predictive performance. More broadly, the ability of models to preserve group-level cognitive differences suggests potential translational value. However, The clinical utility of such models will require prospective validation, evaluation of individual-level reliability, assessment across diverse clinical settings, and consideration of acquisition and computational costs (Arbabshirani et al., 2017).

### Limitations and future directions

Several limitations should be noted. First, ADHD criteria in ABCD are proxy-based rather than derived from full clinical diagnosis, and the external validation cohort was relatively small with limited overlap in available sets of features. These factors constrain broad generalization and motivate future validation in larger, clinically diagnosed ADHD cohorts using harmonized protocols. Second, some major comorbidities, including bipolar disorder, schizophrenia spectrum disorders, or intellectual disability, were addressed through the Tier 2 ADHD definition, but this did not capture all possible comorbidity effects. Future studies could benefit from formally investigating the role of comorbidities during model training and evaluation (Dinga et al., 2020; Weber et al., 2022). Third, the exploratory use of 81 feature sets was intentionally broad rather than based on prespecified key modalities; future studies focused on mechanistic interpretation may benefit from evaluating set of features that contributed most to predictions. Forth, the attenuation of effect size differences between observed and predicted scores in the external dataset further underscores the need for larger samples, greater feature overlap, and harmonized cognitive measures or latent-variable approaches when evaluating cross-dataset transfer. Fifth, although the residualization sensitivity analyses indicated that medication exposure had a negligible impact on predictive performance, medication status was classified using the first-level categories of the Anatomical Therapeutic Chemical (ATC) system. This approach grouped all ADHD medications within a single category, the nervous system class, one of 14 broad ATC categories. Future studies could leverage ADHD-specific datasets that explicitly differentiate between medication types and treatment regimens. Such analyses may help clarify the extent to which medication status influences population-based brain-cognition prediction, particularly for task-based fMRI and functional connectivity measures (Kay et al., 2025; Lopez et al., 2026).

## CONCLUSION

This study provides evidence that multimodal neuroimaging-based cognitive prediction models trained on large-scale population data can transfer robustly to children with ADHD. By systematically benchmarking 81 MRI-derived sets of features and evaluating both internal and external validation, we provide evidence that (i) predicted scores preserve clinically meaningful group-level differences, (ii) stacked models show similar performance across diagnostic tiers with increased stability, and (iii) multimodal integration offers a dual benefit of improved accuracy and improved robustness. While acknowledging the limitations of these models as a fully-fledged clinical tool, our work signals a significant step towards scalable, brain-based tools for understanding cognitive functioning in clinical populations. These findings should be viewed as a step toward understanding brain-based variation in cognition in ADHD.

## ACKNOWLEDGMENTS

This research was supported by the New Zealand Health Research Council (21/618 and 24/838;), the University of Otago, the Neurological Foundation of New Zealand (2350 PRG) and the Ministry of Business, Innovation and Employment (UOA2421 and RTVU2403). The authors acknowledge the use of New Zealand eScience Infrastructure (NeSI) high-performance computing facilities and support services, funded by NeSI collaborator institutions and the Ministry of Business, Innovation & Employment.

This work used publicly available data from two sources. The Adolescent Brain Cognitive Development (ABCD) Study®, supported by the National Institutes of Health and federal partners, provided large-scale, multimodal neuroimaging and cognitive data. A full list of grant numbers is available at https://abcdstudy.org/federal-partners.html, and a list of participating institutions is available at https://abcdstudy.org/consortium_members. ABCD data were accessed via NIMH Data Archive under DOIs: http://dx.doi.org/10.15154/z563-zd24 and http://dx.doi.org/10.15154/1523041.

The second dataset, Working Memory and Reward in Children With and Without Attention-Deficit/Hyperactivity Disorder, was originally published by Lytle and colleagues and is accessible on OpenNeuro (https://openneuro.org/datasets/ds002424/versions/1.2.0).

## Author contributions are as follows

Khakpoor and Pat conceived and designed the study. Data acquisition, analysis, and interpretation were conducted by Khakpoor, Pat, Wang, van der Vliet, and Tetereva. Khakpoor and Pat drafted the manuscript and performed statistical analyses. All authors critically revised the manuscript. Funding was acquired by Pat. Pat also supervised the project. All authors approved the final version and are accountable for the accuracy and integrity of the work.

The authors thank Michael Demidenko (Stanford University) for providing fMRI task event extraction scripts. This manuscript reflects the authors’ views and does not necessarily represent those of any funding body or the ABCD Consortium.

## Data Availability

ABCD Study data are accessible via https://nda.nih.gov/abcd upon NDA approval. The Lytle dataset is available at https://openneuro.org/datasets/ds002424. All analysis code is available at https://github.com/HAM-lab-Otago-University/ADHD-MultiModal.

