## Supplementary Tables for "Predicting Cognitive Functioning in ADHD Using Population-Based MRI Across Large and Small Samples"

|  | **Internal Validation Model** | **External Validation Sets of Features** |
| --- | --- | --- |
| **Single** | Nback 2back | Spatial Nback 2back |
|  | Nback 2back | Verbal Nback 2back |
|  | Nback FC | Verbal Nback FC |
|  | Surface area | Surface area |
|  | General FC | Multitask FC |
|  | Nback 2-0back | Spatial Nback 2-1back |
|  | Nback 0back | Spatial Nback 1back |
|  | Multitask FC | Multitask FC |
|  | Subcortical volume | Subcortical volume |
|  | FreeSurfer summations | FreeSurfer summations |
|  | Nback 2-0back | Verbal Nback 2-1back |
|  | Cortical thickness | Cortical thickness |
|  | Nback FC | Spatial Nback FC |
| **Stacked** | Nback WM load Contrasts | Spatial Nback Contrasts |
|  | ABCC sMRI + Nback FC + Nback WM load Contrasts | sMRI + Verbal Nback FC + Contrasts |
|  | ABCC sMRI + General FC + Nback WM load Contrasts | sMRI + Multitask FC + Spatial Nback Contrasts |
|  | ABCC sMRI + General FC + Nback WM load Contrasts | sMRI + Multitask FC + Verbal Nback Contrasts |
|  | ABCC sMRI + Nback FC + Nback WM load Contrasts | sMRI + Spatial Nback FC + Contrasts |
|  | ABCC sMRI + Multitask FC + Nback WM load Contrasts | sMRI + Multitask FC + Verbal Nback Contrasts |
|  | ABCC sMRI + Multitask FC + Nback WM load Contrasts | sMRI + Multitask FC + Spatial Nback Contrasts |
|  | Nback WM load Contrasts | Verbal Nback Contrasts |
|  | ABCC sMRI | sMRI |

**Supplementary Table 1.** The mapping between models trained in internal validation and the feature sets from external validation.

|  | Original r | Residualised r | Original r | Residualised r | Original r | Residualised r | Original r | Residualised r | Original r | Residualised r | Original r | Residualised r |
| --- | --- | --- | --- | --- | --- | --- | --- | --- | --- | --- | --- | --- |
|  | **All** | | **ADHD -Tier 1** | | **ADHD -Tier 2** | | **ADHD -Tier 3** | | **ADHD -Tier 4** | | **Non-ADHD** | |
| Income | 0.57 | 0.47 | 0.55 | 0.48 | 0.56 | 0.49 | 0.55 | 0.50 | 0.51 | 0.47 | 0.57 | 0.47 |
| Parental education | 0.57 | 0.51 | 0.55 | 0.50 | 0.56 | 0.51 | 0.55 | 0.48 | 0.51 | 0.46 | 0.57 | 0.50 |
| Sex | 0.57 | 0.58 | 0.55 | 0.56 | 0.56 | 0.56 | 0.55 | 0.56 | 0.51 | 0.51 | 0.57 | 0.57 |
| Medication | 0.57 | 0.57 | 0.55 | 0.55 | 0.56 | 0.56 | 0.55 | 0.56 | 0.51 | 0.53 | 0.57 | 0.57 |

**Supplementary Table 2.** Changes in Pearson’s *r* for Stacked All model (best performing model) between observed and predicted cognitive scores before and after residualizing each confounding variable.

|  | **Tier1** | | | | **Tier2** | | | | **Tier3** | | | | **Tier4** | | | | **non-ADHD** | | | | **All** | | | |
| --- | --- | --- | --- | --- | --- | --- | --- | --- | --- | --- | --- | --- | --- | --- | --- | --- | --- | --- | --- | --- | --- | --- | --- | --- |
|  | **r** | **R2** | **MAE** | **RMSE** | **r** | **R2** | **MAE** | **RMSE** | **r** | **R2** | **MAE** | **RMSE** | **r** | **R2** | **MAE** | **RMSE** | **r** | **R2** | **MAE** | **RMSE** | **r** | **R2** | **MAE** | **RMSE** |
| **MID FC** | 0.52 | 0.22 | 0.84 | 0.65 | 0.52 | 0.23 | 0.79 | 0.62 | 0.43 | -0.02 | 0.90 | 0.75 | 0.51 | 0.01 | 0.98 | 0.84 | 0.48 | 0.21 | 0.80 | 0.63 | 0.49 | 0.22 | 0.80 | 0.63 |
| **Rest FC** | 0.50 | 0.20 | 0.93 | 0.72 | 0.50 | 0.21 | 0.89 | 0.70 | 0.47 | -0.02 | 0.85 | 0.72 | 0.58 | 0.15 | 0.80 | 0.68 | 0.44 | 0.18 | 0.84 | 0.67 | 0.45 | 0.19 | 0.85 | 0.68 |
| **General FC** | 0.52 | 0.23 | 0.90 | 0.70 | 0.52 | 0.24 | 0.85 | 0.67 | 0.48 | 0.05 | 0.83 | 0.70 | 0.41 | -0.03 | 0.96 | 0.83 | 0.44 | 0.17 | 0.82 | 0.65 | 0.45 | 0.19 | 0.83 | 0.65 |
| **Glasser Nback 2-0back** | 0.48 | 0.20 | 0.83 | 0.63 | 0.48 | 0.20 | 0.82 | 0.61 | 0.28 | 0.03 | 0.85 | 0.73 | 0.35 | 0.08 | 0.87 | 0.76 | 0.44 | 0.20 | 0.77 | 0.61 | 0.45 | 0.20 | 0.78 | 0.61 |
| **Multitask FC** | 0.49 | 0.19 | 0.83 | 0.65 | 0.48 | 0.18 | 0.81 | 0.63 | 0.54 | 0.05 | 0.80 | 0.65 | 0.44 | -0.06 | 0.91 | 0.76 | 0.44 | 0.18 | 0.80 | 0.63 | 0.45 | 0.18 | 0.80 | 0.63 |
| **Glasser Nback 2back** | 0.43 | 0.15 | 0.86 | 0.65 | 0.42 | 0.15 | 0.84 | 0.64 | 0.50 | 0.19 | 0.78 | 0.65 | 0.66 | 0.28 | 0.77 | 0.65 | 0.43 | 0.18 | 0.78 | 0.62 | 0.43 | 0.18 | 0.78 | 0.62 |
| **Nback FC** | 0.42 | 0.13 | 0.84 | 0.68 | 0.39 | 0.10 | 0.84 | 0.68 | 0.41 | 0.06 | 0.84 | 0.70 | 0.39 | 0.09 | 0.83 | 0.67 | 0.42 | 0.15 | 0.78 | 0.62 | 0.42 | 0.16 | 0.79 | 0.63 |
| **SST FC** | 0.44 | 0.15 | 0.87 | 0.68 | 0.44 | 0.14 | 0.85 | 0.67 | 0.57 | 0.04 | 0.83 | 0.71 | 0.28 | -0.16 | 0.96 | 0.86 | 0.40 | 0.14 | 0.83 | 0.65 | 0.40 | 0.15 | 0.83 | 0.65 |
| **Destriuex Nback 2-0back** | 0.41 | 0.11 | 0.91 | 0.71 | 0.40 | 0.12 | 0.89 | 0.69 | 0.28 | -0.03 | 0.89 | 0.76 | 0.06 | -0.20 | 0.88 | 0.77 | 0.39 | 0.14 | 0.82 | 0.65 | 0.39 | 0.15 | 0.83 | 0.65 |
| **Glasser Nback Face** | 0.33 | 0.07 | 0.90 | 0.69 | 0.32 | 0.07 | 0.88 | 0.68 | 0.51 | 0.16 | 0.79 | 0.67 | 0.63 | 0.20 | 0.81 | 0.69 | 0.38 | 0.14 | 0.80 | 0.64 | 0.38 | 0.14 | 0.81 | 0.64 |
| **Glasser Nback EmotionFace** | 0.32 | 0.06 | 0.90 | 0.69 | 0.31 | 0.07 | 0.88 | 0.68 | 0.54 | 0.15 | 0.80 | 0.70 | 0.61 | 0.16 | 0.83 | 0.73 | 0.37 | 0.14 | 0.80 | 0.64 | 0.37 | 0.14 | 0.81 | 0.64 |
| **Glasser MID Reward-Neutral** | 0.38 | 0.07 | 0.96 | 0.75 | 0.36 | 0.08 | 0.91 | 0.72 | 0.10 | -0.18 | 1.05 | 0.88 | 0.01 | -0.20 | 1.05 | 0.91 | 0.35 | 0.12 | 0.86 | 0.68 | 0.36 | 0.13 | 0.86 | 0.68 |
| **Glasser Nback 0back** | 0.32 | 0.06 | 0.90 | 0.69 | 0.32 | 0.07 | 0.88 | 0.68 | 0.33 | 0.03 | 0.85 | 0.73 | 0.26 | 0.00 | 0.90 | 0.80 | 0.35 | 0.12 | 0.81 | 0.64 | 0.35 | 0.12 | 0.81 | 0.65 |
| **Destriuex Nback 2back** | 0.32 | 0.06 | 0.94 | 0.73 | 0.30 | 0.06 | 0.92 | 0.72 | 0.20 | -0.07 | 0.90 | 0.75 | 0.12 | -0.14 | 0.86 | 0.73 | 0.35 | 0.11 | 0.83 | 0.66 | 0.35 | 0.11 | 0.84 | 0.66 |
| **Glasser MID LargeReward-Neutral** | 0.37 | 0.07 | 0.97 | 0.76 | 0.34 | 0.07 | 0.92 | 0.73 | 0.04 | -0.20 | 1.06 | 0.90 | -0.06 | -0.26 | 1.08 | 0.92 | 0.34 | 0.11 | 0.86 | 0.68 | 0.35 | 0.12 | 0.87 | 0.69 |
| **Glasser Nback Place** | 0.35 | 0.07 | 0.90 | 0.69 | 0.35 | 0.08 | 0.87 | 0.68 | 0.50 | 0.11 | 0.81 | 0.67 | 0.61 | 0.15 | 0.83 | 0.70 | 0.34 | 0.11 | 0.81 | 0.64 | 0.34 | 0.12 | 0.82 | 0.65 |
| **Glasser MID Loss-Neutral** | 0.38 | 0.06 | 0.97 | 0.76 | 0.37 | 0.07 | 0.92 | 0.72 | 0.24 | -0.13 | 1.03 | 0.87 | 0.15 | -0.13 | 1.03 | 0.91 | 0.33 | 0.11 | 0.86 | 0.68 | 0.34 | 0.11 | 0.87 | 0.69 |
| **Glasser MID LargeLoss-Neutral** | 0.39 | 0.06 | 0.97 | 0.76 | 0.38 | 0.07 | 0.92 | 0.72 | 0.29 | -0.12 | 1.03 | 0.86 | 0.21 | -0.12 | 1.02 | 0.90 | 0.32 | 0.10 | 0.87 | 0.69 | 0.33 | 0.11 | 0.87 | 0.69 |
| **Glasser MID SmallReward-Neutral** | 0.35 | 0.04 | 0.98 | 0.76 | 0.34 | 0.06 | 0.92 | 0.72 | 0.13 | -0.18 | 1.05 | 0.86 | 0.15 | -0.11 | 1.01 | 0.87 | 0.32 | 0.10 | 0.86 | 0.68 | 0.33 | 0.11 | 0.87 | 0.69 |
| **Glasser MID SmallLoss-Neutral** | 0.34 | 0.03 | 0.99 | 0.77 | 0.32 | 0.04 | 0.93 | 0.73 | 0.08 | -0.19 | 1.06 | 0.89 | 0.02 | -0.18 | 1.05 | 0.93 | 0.31 | 0.10 | 0.87 | 0.69 | 0.32 | 0.10 | 0.88 | 0.69 |
| **rs-fMRI Temporal Variance** | 0.23 | -0.03 | 1.06 | 0.82 | 0.22 | -0.03 | 1.01 | 0.79 | 0.14 | -0.53 | 1.28 | 1.01 | 0.12 | -0.91 | 1.28 | 0.97 | 0.32 | 0.10 | 0.91 | 0.72 | 0.31 | 0.10 | 0.92 | 0.73 |
| **Glasser MID LossHit-LossMiss** | 0.30 | -0.01 | 1.00 | 0.78 | 0.29 | 0.01 | 0.94 | 0.75 | 0.28 | -0.15 | 1.04 | 0.86 | 0.27 | -0.13 | 1.03 | 0.87 | 0.30 | 0.09 | 0.87 | 0.69 | 0.30 | 0.09 | 0.88 | 0.70 |
| **Glasser MID RewardHit-RewardMiss** | 0.33 | 0.02 | 0.99 | 0.77 | 0.33 | 0.04 | 0.93 | 0.73 | 0.19 | -0.10 | 1.02 | 0.86 | -0.08 | -0.12 | 1.02 | 0.90 | 0.29 | 0.09 | 0.87 | 0.69 | 0.30 | 0.09 | 0.88 | 0.70 |
| **rsfMRI Subcortical-network FC** | 0.34 | 0.03 | 1.03 | 0.81 | 0.32 | 0.04 | 0.98 | 0.78 | 0.37 | -0.15 | 1.11 | 0.90 | 0.31 | -0.23 | 1.03 | 0.84 | 0.30 | 0.09 | 0.92 | 0.73 | 0.30 | 0.09 | 0.93 | 0.73 |
| **rsfMRI cortical FC** | 0.29 | -0.01 | 1.05 | 0.82 | 0.30 | 0.02 | 0.99 | 0.79 | 0.24 | -0.25 | 1.16 | 0.94 | 0.29 | -0.27 | 1.04 | 0.86 | 0.30 | 0.08 | 0.92 | 0.73 | 0.30 | 0.08 | 0.93 | 0.73 |
| **Cortical Thickness** | 0.28 | -0.01 | 1.07 | 0.83 | 0.29 | 0.02 | 1.02 | 0.81 | 0.28 | -0.25 | 1.17 | 0.94 | 0.29 | -0.25 | 1.10 | 0.89 | 0.28 | 0.07 | 0.95 | 0.75 | 0.28 | 0.08 | 0.96 | 0.76 |
| **T2 Gray Matter Avg Intensity** | 0.28 | 0.00 | 1.05 | 0.82 | 0.27 | 0.02 | 0.99 | 0.79 | 0.27 | -0.25 | 1.09 | 0.88 | 0.26 | -0.27 | 0.98 | 0.80 | 0.28 | 0.07 | 0.94 | 0.74 | 0.28 | 0.07 | 0.95 | 0.75 |
| **Glasser SST Stop-CorrectGo** | 0.24 | -0.01 | 0.99 | 0.77 | 0.24 | 0.00 | 0.96 | 0.75 | 0.29 | -0.31 | 0.98 | 0.81 | 0.17 | -0.42 | 1.07 | 0.91 | 0.28 | 0.08 | 0.87 | 0.69 | 0.28 | 0.08 | 0.88 | 0.69 |
| **Glasser SST IncorrectStop** | 0.19 | -0.03 | 1.00 | 0.78 | 0.20 | -0.02 | 0.97 | 0.76 | 0.24 | -0.34 | 1.00 | 0.85 | 0.10 | -0.45 | 1.08 | 0.95 | 0.28 | 0.07 | 0.87 | 0.69 | 0.27 | 0.07 | 0.88 | 0.70 |
| **Surface Area** | 0.26 | -0.04 | 1.05 | 0.83 | 0.26 | -0.03 | 1.02 | 0.81 | 0.26 | -0.35 | 1.17 | 0.95 | 0.41 | -0.17 | 1.10 | 0.90 | 0.27 | 0.07 | 0.93 | 0.73 | 0.27 | 0.07 | 0.94 | 0.74 |
| **Glasser SST IncorrectGo** | 0.21 | -0.02 | 0.99 | 0.78 | 0.22 | -0.01 | 0.96 | 0.76 | 0.29 | -0.26 | 0.96 | 0.81 | -0.06 | -0.39 | 1.06 | 0.92 | 0.26 | 0.07 | 0.87 | 0.69 | 0.26 | 0.07 | 0.88 | 0.70 |
| **T1 Gray Matter Avg Intensity** | 0.25 | -0.01 | 1.07 | 0.84 | 0.26 | 0.01 | 1.03 | 0.81 | 0.34 | -0.20 | 1.15 | 0.92 | 0.35 | -0.20 | 1.08 | 0.88 | 0.26 | 0.07 | 0.95 | 0.75 | 0.26 | 0.07 | 0.96 | 0.76 |
| **Glasser SST IncorrectStop-CorrectGo** | 0.21 | -0.03 | 1.00 | 0.78 | 0.21 | -0.02 | 0.97 | 0.76 | 0.19 | -0.39 | 1.01 | 0.84 | 0.15 | -0.48 | 1.09 | 0.94 | 0.26 | 0.07 | 0.87 | 0.69 | 0.26 | 0.07 | 0.88 | 0.70 |
| **Glasser SST IncorrectGo-CorrectGo** | 0.25 | 0.00 | 0.99 | 0.77 | 0.25 | 0.01 | 0.96 | 0.75 | 0.46 | -0.14 | 0.92 | 0.78 | 0.35 | -0.24 | 1.00 | 0.89 | 0.26 | 0.07 | 0.87 | 0.69 | 0.26 | 0.07 | 0.88 | 0.70 |
| **Glasser SST CorrectStop** | 0.21 | -0.02 | 1.00 | 0.78 | 0.21 | -0.01 | 0.96 | 0.76 | 0.30 | -0.32 | 0.99 | 0.84 | 0.06 | -0.41 | 1.07 | 0.94 | 0.26 | 0.06 | 0.87 | 0.69 | 0.26 | 0.06 | 0.88 | 0.70 |
| **T2 White Matter Avg Intensity** | 0.24 | -0.01 | 1.05 | 0.83 | 0.24 | 0.01 | 1.00 | 0.80 | 0.26 | -0.24 | 1.08 | 0.88 | 0.27 | -0.25 | 0.97 | 0.80 | 0.25 | 0.06 | 0.95 | 0.75 | 0.25 | 0.06 | 0.96 | 0.75 |
| **Destriuex Nback Emotionface** | 0.19 | -0.02 | 0.98 | 0.76 | 0.18 | -0.01 | 0.96 | 0.74 | 0.27 | -0.10 | 0.92 | 0.79 | 0.20 | -0.21 | 0.89 | 0.78 | 0.26 | 0.06 | 0.86 | 0.68 | 0.25 | 0.06 | 0.87 | 0.68 |
| **Destriuex Nback 0back** | 0.20 | -0.02 | 0.98 | 0.76 | 0.21 | 0.00 | 0.95 | 0.75 | 0.38 | -0.09 | 0.91 | 0.79 | 0.14 | -0.28 | 0.91 | 0.81 | 0.26 | 0.06 | 0.86 | 0.68 | 0.25 | 0.06 | 0.87 | 0.68 |
| **Cortical Volume** | 0.29 | -0.03 | 1.08 | 0.85 | 0.29 | 0.00 | 1.04 | 0.82 | 0.29 | -0.31 | 1.20 | 0.96 | 0.33 | -0.30 | 1.13 | 0.91 | 0.25 | 0.06 | 0.96 | 0.76 | 0.25 | 0.06 | 0.97 | 0.77 |
| **T1 Normalised Intensity** | 0.24 | -0.02 | 1.07 | 0.84 | 0.25 | 0.01 | 1.03 | 0.82 | 0.32 | -0.21 | 1.15 | 0.94 | 0.31 | -0.22 | 1.09 | 0.90 | 0.25 | 0.06 | 0.96 | 0.76 | 0.25 | 0.06 | 0.97 | 0.77 |
| **T1 White Matter Avg Intensity** | 0.24 | -0.03 | 1.08 | 0.84 | 0.25 | 0.00 | 1.03 | 0.82 | 0.25 | -0.27 | 1.18 | 0.94 | 0.36 | -0.22 | 1.09 | 0.89 | 0.24 | 0.06 | 0.96 | 0.76 | 0.24 | 0.06 | 0.97 | 0.77 |
| **T2 Normalised Intensity** | 0.23 | -0.02 | 1.06 | 0.82 | 0.24 | 0.00 | 1.00 | 0.79 | 0.09 | -0.35 | 1.13 | 0.90 | 0.04 | -0.38 | 1.02 | 0.83 | 0.24 | 0.05 | 0.95 | 0.75 | 0.24 | 0.06 | 0.96 | 0.76 |
| **Glasser SST CorrectStop-CorrectGo** | 0.23 | -0.01 | 0.99 | 0.77 | 0.26 | 0.00 | 0.96 | 0.75 | 0.38 | -0.26 | 0.97 | 0.79 | 0.46 | -0.31 | 1.03 | 0.87 | 0.24 | 0.06 | 0.88 | 0.70 | 0.24 | 0.06 | 0.89 | 0.70 |
| **Destriuex MID SmallReward-Neutral** | 0.26 | -0.02 | 1.05 | 0.82 | 0.26 | 0.00 | 1.00 | 0.79 | 0.19 | -0.27 | 1.06 | 0.87 | 0.16 | -0.32 | 1.00 | 0.83 | 0.23 | 0.05 | 0.93 | 0.74 | 0.23 | 0.05 | 0.94 | 0.74 |
| **Glasser MID LargeReward-SmallReward** | 0.23 | -0.03 | 1.01 | 0.80 | 0.21 | -0.01 | 0.96 | 0.76 | 0.02 | -0.20 | 1.06 | 0.87 | -0.05 | -0.21 | 1.06 | 0.89 | 0.23 | 0.05 | 0.89 | 0.70 | 0.23 | 0.05 | 0.90 | 0.71 |
| **Glasser SST CorrectGo** | 0.16 | -0.04 | 1.01 | 0.79 | 0.17 | -0.03 | 0.98 | 0.77 | 0.20 | -0.37 | 1.01 | 0.89 | -0.04 | -0.47 | 1.09 | 0.99 | 0.23 | 0.05 | 0.88 | 0.70 | 0.23 | 0.05 | 0.89 | 0.70 |
| **T1 Summations** | 0.21 | -0.06 | 1.09 | 0.85 | 0.22 | -0.03 | 1.05 | 0.83 | 0.17 | -0.33 | 1.21 | 0.97 | 0.21 | -0.32 | 1.14 | 0.91 | 0.23 | 0.05 | 0.96 | 0.76 | 0.23 | 0.05 | 0.97 | 0.77 |
| **Destriuex MID Loss-Neutral** | 0.24 | -0.02 | 1.05 | 0.82 | 0.23 | -0.01 | 1.00 | 0.79 | 0.20 | -0.26 | 1.05 | 0.87 | 0.16 | -0.30 | 0.99 | 0.83 | 0.22 | 0.04 | 0.93 | 0.74 | 0.22 | 0.05 | 0.94 | 0.74 |
| **Destriuex MID Reward-Neutral** | 0.27 | -0.02 | 1.05 | 0.82 | 0.25 | 0.00 | 1.00 | 0.79 | 0.25 | -0.26 | 1.05 | 0.86 | 0.23 | -0.31 | 0.99 | 0.81 | 0.22 | 0.04 | 0.93 | 0.74 | 0.22 | 0.04 | 0.94 | 0.74 |
| **Glasser Nback Face-Place** | 0.17 | -0.03 | 0.95 | 0.72 | 0.18 | -0.01 | 0.92 | 0.70 | 0.36 | -0.06 | 0.89 | 0.72 | 0.44 | -0.04 | 0.92 | 0.78 | 0.23 | 0.05 | 0.84 | 0.67 | 0.22 | 0.05 | 0.85 | 0.67 |
| **Glasser SST IncorrectGo-IncorrectStop** | 0.17 | -0.04 | 1.01 | 0.79 | 0.17 | -0.04 | 0.98 | 0.77 | 0.45 | -0.22 | 0.95 | 0.79 | 0.35 | -0.29 | 1.02 | 0.88 | 0.22 | 0.05 | 0.88 | 0.70 | 0.22 | 0.05 | 0.89 | 0.71 |
| **Destriuex MID LargeReward-Neutral** | 0.25 | -0.02 | 1.05 | 0.82 | 0.23 | 0.00 | 1.00 | 0.79 | 0.06 | -0.31 | 1.07 | 0.88 | 0.05 | -0.36 | 1.01 | 0.83 | 0.21 | 0.04 | 0.93 | 0.74 | 0.22 | 0.04 | 0.94 | 0.74 |
| **Destriuex Nback Place** | 0.17 | -0.03 | 0.98 | 0.77 | 0.17 | -0.02 | 0.96 | 0.75 | 0.13 | -0.18 | 0.95 | 0.81 | -0.01 | -0.32 | 0.92 | 0.80 | 0.21 | 0.04 | 0.87 | 0.68 | 0.21 | 0.04 | 0.88 | 0.69 |
| **FreeSurfer summations** | 0.22 | -0.07 | 1.07 | 0.84 | 0.22 | -0.05 | 1.03 | 0.82 | 0.16 | -0.40 | 1.19 | 0.97 | 0.34 | -0.26 | 1.14 | 0.93 | 0.21 | 0.04 | 0.95 | 0.75 | 0.21 | 0.04 | 0.96 | 0.76 |
| **Glasser MID LargeLoss-SmallLoss** | 0.24 | -0.04 | 1.02 | 0.79 | 0.24 | -0.02 | 0.96 | 0.75 | -0.01 | -0.27 | 1.09 | 0.90 | 0.01 | -0.21 | 1.06 | 0.92 | 0.20 | 0.04 | 0.90 | 0.71 | 0.20 | 0.04 | 0.91 | 0.71 |
| **Destriuex MID SmallLoss-Neutral** | 0.24 | -0.04 | 1.06 | 0.83 | 0.21 | -0.02 | 1.01 | 0.80 | 0.24 | -0.31 | 1.07 | 0.87 | 0.27 | -0.33 | 1.00 | 0.82 | 0.19 | 0.03 | 0.94 | 0.74 | 0.20 | 0.03 | 0.95 | 0.75 |
| **Destriuex SST AnyStop-CorrectStop** | 0.15 | -0.05 | 1.06 | 0.83 | 0.14 | -0.04 | 1.02 | 0.81 | 0.10 | -0.42 | 1.03 | 0.87 | 0.18 | -0.67 | 1.02 | 0.87 | 0.19 | 0.03 | 0.92 | 0.72 | 0.19 | 0.03 | 0.93 | 0.73 |
| **Destriuex MID LossHit-LossMiss** | 0.20 | -0.04 | 1.06 | 0.83 | 0.19 | -0.02 | 1.01 | 0.80 | 0.16 | -0.30 | 1.07 | 0.87 | 0.13 | -0.35 | 1.01 | 0.82 | 0.18 | 0.03 | 0.94 | 0.74 | 0.19 | 0.03 | 0.95 | 0.75 |
| **Destriuex MID LargeLoss-Neutral** | 0.24 | -0.04 | 1.06 | 0.82 | 0.21 | -0.02 | 1.01 | 0.79 | 0.21 | -0.29 | 1.06 | 0.87 | 0.08 | -0.35 | 1.01 | 0.83 | 0.17 | 0.03 | 0.94 | 0.74 | 0.18 | 0.03 | 0.95 | 0.75 |
| **Glasser SST CorrectStop-IncorrectStop** | 0.08 | -0.07 | 1.02 | 0.80 | 0.09 | -0.06 | 0.99 | 0.78 | 0.10 | -0.41 | 1.02 | 0.87 | -0.01 | -0.48 | 1.10 | 0.96 | 0.18 | 0.03 | 0.89 | 0.70 | 0.17 | 0.03 | 0.90 | 0.71 |
| **Destriuex SST CorrectStop-CorrectGo** | 0.13 | -0.06 | 1.06 | 0.83 | 0.12 | -0.04 | 1.02 | 0.81 | 0.17 | -0.39 | 1.02 | 0.86 | 0.22 | -0.63 | 1.01 | 0.86 | 0.18 | 0.03 | 0.92 | 0.73 | 0.17 | 0.03 | 0.93 | 0.73 |
| **Destriuex SST IncorrectStop-CorrectGo** | 0.13 | -0.06 | 1.06 | 0.83 | 0.12 | -0.05 | 1.02 | 0.81 | -0.01 | -0.47 | 1.04 | 0.88 | 0.03 | -0.74 | 1.04 | 0.88 | 0.17 | 0.03 | 0.92 | 0.73 | 0.17 | 0.03 | 0.93 | 0.73 |
| **Destriuex MID RewardHit-RewardMiss** | 0.16 | -0.05 | 1.06 | 0.83 | 0.18 | -0.02 | 1.01 | 0.80 | 0.20 | -0.28 | 1.06 | 0.85 | 0.14 | -0.37 | 1.01 | 0.84 | 0.17 | 0.02 | 0.94 | 0.74 | 0.17 | 0.03 | 0.95 | 0.75 |
| **T2 Subcortical Volume** | 0.12 | -0.06 | 1.08 | 0.85 | 0.13 | -0.04 | 1.02 | 0.81 | 0.13 | -0.34 | 1.12 | 0.91 | 0.03 | -0.39 | 1.03 | 0.83 | 0.16 | 0.02 | 0.97 | 0.76 | 0.16 | 0.02 | 0.98 | 0.77 |
| **T1 Subcortical Volume** | 0.14 | -0.07 | 1.10 | 0.86 | 0.16 | -0.04 | 1.05 | 0.83 | 0.20 | -0.33 | 1.21 | 0.97 | 0.15 | -0.37 | 1.16 | 0.94 | 0.16 | 0.02 | 0.97 | 0.77 | 0.16 | 0.03 | 0.99 | 0.78 |
| **Destriuex MID LargeReward-SmallReward** | 0.17 | -0.05 | 1.06 | 0.84 | 0.18 | -0.03 | 1.01 | 0.81 | 0.05 | -0.33 | 1.08 | 0.89 | 0.01 | -0.39 | 1.02 | 0.84 | 0.15 | 0.02 | 0.94 | 0.75 | 0.15 | 0.02 | 0.95 | 0.75 |
| **DTI** | 0.12 | -0.06 | 1.08 | 0.85 | 0.11 | -0.04 | 1.03 | 0.82 | 0.14 | -0.34 | 1.15 | 0.91 | 0.19 | -0.36 | 1.04 | 0.84 | 0.14 | 0.02 | 0.97 | 0.76 | 0.13 | 0.02 | 0.98 | 0.77 |
| **Destriuex SST CorrectGo-Fixation** | 0.07 | -0.07 | 1.06 | 0.83 | 0.05 | -0.06 | 1.03 | 0.81 | 0.08 | -0.42 | 1.03 | 0.87 | 0.02 | -0.69 | 1.03 | 0.89 | 0.13 | 0.02 | 0.92 | 0.73 | 0.12 | 0.01 | 0.93 | 0.74 |
| **Destriuex Nback Face-Place** | 0.12 | -0.05 | 0.99 | 0.77 | 0.11 | -0.04 | 0.97 | 0.75 | 0.04 | -0.22 | 0.97 | 0.81 | 0.05 | -0.36 | 0.94 | 0.81 | 0.12 | 0.01 | 0.88 | 0.70 | 0.12 | 0.01 | 0.89 | 0.70 |
| **Destriuex MID LargeLoss-SmallLoss** | 0.07 | -0.08 | 1.08 | 0.84 | 0.05 | -0.06 | 1.03 | 0.81 | 0.14 | -0.32 | 1.08 | 0.87 | 0.07 | -0.39 | 1.02 | 0.83 | 0.11 | 0.01 | 0.95 | 0.75 | 0.11 | 0.01 | 0.96 | 0.75 |
| **Glasser Nback NegFace-NeutFace** | 0.04 | -0.06 | 0.96 | 0.73 | 0.04 | -0.06 | 0.94 | 0.71 | -0.04 | -0.21 | 0.95 | 0.78 | 0.06 | -0.21 | 1.00 | 0.86 | 0.12 | 0.01 | 0.85 | 0.68 | 0.11 | 0.01 | 0.86 | 0.68 |
| **Destriuex SST CorrectStop-IncorrectStop** | 0.08 | -0.07 | 1.06 | 0.83 | 0.10 | -0.05 | 1.03 | 0.81 | 0.08 | -0.46 | 1.04 | 0.87 | 0.01 | -0.74 | 1.04 | 0.88 | 0.11 | 0.01 | 0.93 | 0.73 | 0.11 | 0.01 | 0.94 | 0.74 |
| **Destriuex SST IncorrectGo-CorrectGo** | 0.13 | -0.06 | 1.06 | 0.83 | 0.13 | -0.04 | 1.02 | 0.81 | 0.04 | -0.41 | 1.02 | 0.87 | 0.12 | -0.66 | 1.02 | 0.88 | 0.10 | 0.01 | 0.93 | 0.73 | 0.10 | 0.01 | 0.94 | 0.74 |
| **Glasser Nback Emotionface-Neutralface** | 0.03 | -0.07 | 0.96 | 0.73 | 0.03 | -0.06 | 0.94 | 0.72 | -0.03 | -0.20 | 0.94 | 0.79 | -0.10 | -0.23 | 1.00 | 0.87 | 0.11 | 0.01 | 0.86 | 0.68 | 0.10 | 0.01 | 0.86 | 0.69 |
| **Destriuex SST IncorrectGo-IncorrectStop** | 0.02 | -0.07 | 1.07 | 0.84 | 0.04 | -0.06 | 1.03 | 0.81 | -0.06 | -0.43 | 1.03 | 0.88 | 0.00 | -0.68 | 1.02 | 0.89 | 0.07 | 0.00 | 0.93 | 0.73 | 0.06 | 0.00 | 0.94 | 0.74 |
| **T2 Summations** | 0.05 | -0.08 | 1.09 | 0.85 | 0.05 | -0.06 | 1.03 | 0.82 | -0.09 | -0.39 | 1.14 | 0.93 | -0.24 | -0.41 | 1.03 | 0.85 | 0.05 | 0.00 | 0.98 | 0.77 | 0.05 | 0.00 | 0.99 | 0.78 |
| **Destriuex Nback Emotionface-Neutralface** | 0.04 | -0.06 | 1.00 | 0.77 | 0.03 | -0.05 | 0.97 | 0.76 | 0.08 | -0.21 | 0.96 | 0.82 | 0.02 | -0.35 | 0.93 | 0.83 | 0.03 | 0.00 | 0.88 | 0.70 | 0.03 | 0.00 | 0.89 | 0.71 |
| **Destriuex Nback PosFace-NeutFace** | 0.04 | -0.06 | 1.00 | 0.77 | 0.03 | -0.05 | 0.98 | 0.76 | 0.09 | -0.21 | 0.96 | 0.82 | 0.01 | -0.35 | 0.93 | 0.83 | 0.03 | 0.00 | 0.88 | 0.70 | 0.03 | 0.00 | 0.89 | 0.71 |
| **Destriuex Nback NegFace-NeutFace** | -0.02 | -0.07 | 1.00 | 0.77 | -0.03 | -0.05 | 0.98 | 0.76 | 0.06 | -0.21 | 0.97 | 0.82 | 0.11 | -0.35 | 0.93 | 0.83 | 0.03 | 0.00 | 0.89 | 0.70 | 0.03 | 0.00 | 0.89 | 0.71 |
| **Glasser Nback Posface-Neutralface** | -0.05 | -0.07 | 0.96 | 0.73 | -0.05 | -0.06 | 0.94 | 0.72 | -0.02 | -0.18 | 0.94 | 0.77 | -0.29 | -0.22 | 1.00 | 0.85 | 0.03 | 0.00 | 0.86 | 0.69 | 0.02 | 0.00 | 0.87 | 0.69 |
| **Sulcal Depth** | 0.06 | -0.10 | 1.11 | 0.87 | 0.06 | -0.07 | 1.07 | 0.85 | -0.09 | -0.39 | 1.24 | 1.00 | -0.04 | -0.41 | 1.17 | 0.96 | 0.01 | 0.00 | 0.99 | 0.78 | 0.02 | 0.00 | 1.00 | 0.79 |

**Supplementary Table 3.** Performance metrics of first layer single modalities across non-adhd and adhd tiers in internal validation.

|  | **Tier1** | | | | **Tier2** | | | | **Tier3** | | | | **Tier4** | | | | **non-ADHD** | | | | **All** | | | |
| --- | --- | --- | --- | --- | --- | --- | --- | --- | --- | --- | --- | --- | --- | --- | --- | --- | --- | --- | --- | --- | --- | --- | --- | --- |
|  | **r** | **R2** | **MAE** | **RMSE** | **r** | **R2** | **MAE** | **RMSE** | **r** | **R2** | **MAE** | **RMSE** | **r** | **R2** | **MAE** | **RMSE** | **r** | **R2** | **MAE** | **RMSE** | **r** | **R2** | **MAE** | **RMSE** |
| **Stacked All** | 0.55 | 0.29 | 0.90 | 0.70 | 0.56 | 0.29 | 0.87 | 0.69 | 0.56 | 0.21 | 0.93 | 0.76 | 0.52 | 0.17 | 0.89 | 0.74 | 0.57 | 0.33 | 0.81 | 0.64 | 0.57 | 0.33 | 0.82 | 0.64 |
| **Task FCs + Contrasts** | 0.55 | 0.28 | 0.88 | 0.68 | 0.54 | 0.28 | 0.85 | 0.67 | 0.48 | 0.16 | 0.86 | 0.70 | 0.34 | 0.05 | 0.86 | 0.71 | 0.56 | 0.31 | 0.79 | 0.63 | 0.56 | 0.32 | 0.80 | 0.63 |
| **Task Contrasts** | 0.55 | 0.28 | 0.87 | 0.68 | 0.54 | 0.28 | 0.84 | 0.66 | 0.49 | 0.19 | 0.85 | 0.70 | 0.38 | 0.12 | 0.83 | 0.70 | 0.55 | 0.30 | 0.80 | 0.63 | 0.55 | 0.30 | 0.81 | 0.64 |
| **ABCC Task and Rest FCs** | 0.54 | 0.26 | 0.86 | 0.68 | 0.54 | 0.27 | 0.83 | 0.66 | 0.55 | 0.15 | 0.83 | 0.68 | 0.54 | 0.17 | 0.84 | 0.71 | 0.53 | 0.28 | 0.79 | 0.63 | 0.53 | 0.28 | 0.80 | 0.63 |
| **All FCs** | 0.53 | 0.25 | 0.87 | 0.68 | 0.53 | 0.26 | 0.83 | 0.66 | 0.54 | 0.13 | 0.84 | 0.70 | 0.53 | 0.15 | 0.85 | 0.72 | 0.53 | 0.27 | 0.79 | 0.63 | 0.53 | 0.27 | 0.80 | 0.63 |
| **Task FCs + Contrasts** | 0.51 | 0.23 | 0.85 | 0.67 | 0.51 | 0.23 | 0.83 | 0.66 | 0.49 | 0.10 | 0.86 | 0.71 | 0.46 | 0.08 | 0.94 | 0.81 | 0.52 | 0.26 | 0.79 | 0.62 | 0.52 | 0.26 | 0.79 | 0.63 |
| **ABCC sMRI + Multitask FC + Nback WM Load Contrasts** | 0.51 | 0.22 | 0.92 | 0.70 | 0.51 | 0.22 | 0.89 | 0.69 | 0.41 | 0.03 | 0.99 | 0.78 | 0.49 | 0.16 | 0.92 | 0.73 | 0.50 | 0.25 | 0.84 | 0.66 | 0.51 | 0.25 | 0.85 | 0.66 |
| **ABCC sMRI + General FC + Nback WM Load Contrasts** | 0.51 | 0.23 | 0.91 | 0.70 | 0.51 | 0.23 | 0.88 | 0.69 | 0.40 | 0.04 | 0.98 | 0.78 | 0.47 | 0.14 | 0.93 | 0.73 | 0.50 | 0.25 | 0.84 | 0.66 | 0.51 | 0.25 | 0.85 | 0.66 |
| **ABCC sMRI + Nback FC + Nback WM Load Contrasts** | 0.48 | 0.19 | 0.93 | 0.72 | 0.49 | 0.20 | 0.90 | 0.70 | 0.39 | 0.00 | 1.00 | 0.80 | 0.49 | 0.14 | 0.93 | 0.73 | 0.49 | 0.24 | 0.85 | 0.66 | 0.49 | 0.24 | 0.85 | 0.67 |
| **Nback WM Load Contrasts** | 0.49 | 0.22 | 0.82 | 0.62 | 0.48 | 0.22 | 0.81 | 0.62 | 0.36 | 0.11 | 0.82 | 0.70 | 0.50 | 0.24 | 0.79 | 0.66 | 0.48 | 0.23 | 0.76 | 0.60 | 0.49 | 0.24 | 0.76 | 0.60 |
| **Nback Contrasts + FC** | 0.47 | 0.20 | 0.87 | 0.68 | 0.45 | 0.19 | 0.86 | 0.68 | 0.49 | 0.18 | 0.79 | 0.67 | 0.42 | 0.12 | 0.76 | 0.64 | 0.48 | 0.22 | 0.78 | 0.62 | 0.48 | 0.23 | 0.79 | 0.62 |
| **Nback Contrasts** | 0.45 | 0.19 | 0.88 | 0.68 | 0.44 | 0.18 | 0.86 | 0.67 | 0.42 | 0.14 | 0.81 | 0.68 | 0.36 | 0.08 | 0.77 | 0.65 | 0.47 | 0.22 | 0.78 | 0.62 | 0.48 | 0.23 | 0.79 | 0.62 |
| **Non-Task** | 0.48 | 0.20 | 0.95 | 0.75 | 0.48 | 0.20 | 0.92 | 0.73 | 0.47 | 0.08 | 1.00 | 0.81 | 0.53 | 0.12 | 0.92 | 0.75 | 0.47 | 0.22 | 0.87 | 0.69 | 0.47 | 0.22 | 0.88 | 0.70 |
| **MID Contrasts + FC** | 0.48 | 0.20 | 0.93 | 0.73 | 0.47 | 0.20 | 0.89 | 0.71 | 0.39 | 0.01 | 0.93 | 0.75 | 0.39 | 0.00 | 0.87 | 0.72 | 0.46 | 0.20 | 0.85 | 0.67 | 0.46 | 0.21 | 0.86 | 0.68 |
| **Rest FCs** | 0.47 | 0.17 | 0.95 | 0.75 | 0.45 | 0.17 | 0.92 | 0.73 | 0.46 | 0.08 | 0.99 | 0.83 | 0.48 | 0.05 | 0.90 | 0.75 | 0.44 | 0.19 | 0.87 | 0.69 | 0.44 | 0.19 | 0.87 | 0.69 |
| **MID Contrasts** | 0.46 | 0.17 | 0.95 | 0.75 | 0.45 | 0.17 | 0.91 | 0.72 | 0.40 | -0.01 | 0.94 | 0.76 | 0.34 | -0.06 | 0.89 | 0.74 | 0.41 | 0.16 | 0.87 | 0.69 | 0.41 | 0.17 | 0.88 | 0.69 |
| **sMRI** | 0.40 | 0.10 | 1.01 | 0.79 | 0.41 | 0.13 | 0.97 | 0.77 | 0.41 | -0.03 | 1.06 | 0.84 | 0.47 | 0.02 | 0.97 | 0.78 | 0.39 | 0.15 | 0.91 | 0.72 | 0.40 | 0.16 | 0.92 | 0.72 |
| **SST Contrasts + FC** | 0.34 | 0.07 | 1.00 | 0.78 | 0.34 | 0.07 | 0.97 | 0.76 | 0.49 | -0.09 | 0.90 | 0.72 | 0.24 | -0.41 | 0.94 | 0.79 | 0.38 | 0.14 | 0.86 | 0.68 | 0.38 | 0.14 | 0.87 | 0.69 |
| **ABCC sMRI** | 0.34 | 0.03 | 1.02 | 0.79 | 0.34 | 0.04 | 0.99 | 0.77 | 0.32 | -0.24 | 1.12 | 0.90 | 0.41 | -0.10 | 1.06 | 0.85 | 0.34 | 0.11 | 0.91 | 0.72 | 0.34 | 0.11 | 0.92 | 0.72 |
| **SST Contrasts** | 0.25 | 0.00 | 1.03 | 0.81 | 0.25 | 0.01 | 1.00 | 0.79 | 0.38 | -0.20 | 0.95 | 0.77 | 0.19 | -0.49 | 0.97 | 0.81 | 0.32 | 0.10 | 0.88 | 0.70 | 0.32 | 0.10 | 0.89 | 0.71 |

**Supplementary Table 4.** Performance metrics of second layer stacked modalities across non-adhd and adhd tiers in internal validation.

**Supplementary Table 5.** Performance metrics of first layer stacked modalities across non-adhd and adhd tiers in external validation.

|  | **ADHD** | | | | **non-ADHD** | | | | **All** | | | |
| --- | --- | --- | --- | --- | --- | --- | --- | --- | --- | --- | --- | --- |
|  | **r** | **R2** | **MAE** | **RMSE** | **r** | **R2** | **MAE** | **RMSE** | **r** | **R2** | **MAE** | **RMSE** |
| **Spatial Nback 2back** | 0.31 | -0.27 | 1.04 | 0.88 | 0.24 | 0.05 | 0.95 | 0.79 | 0.38 | 0.09 | 0.99 | 0.83 |
| **Verbal Nback 2back** | 0.27 | -0.39 | 1.01 | 0.82 | 0.26 | 0.06 | 0.93 | 0.78 | 0.35 | 0.08 | 0.96 | 0.80 |
| **Verbal Nback FC** | 0.08 | -0.64 | 1.16 | 0.92 | 0.42 | 0.13 | 0.93 | 0.79 | 0.35 | 0.05 | 1.02 | 0.83 |
| **Surface area** | 0.35 | -0.19 | 0.94 | 0.76 | 0.30 | -0.02 | 0.97 | 0.81 | 0.34 | 0.10 | 0.95 | 0.79 |
| **G-Multitask FC** | -0.02 | -0.41 | 1.11 | 0.86 | 0.48 | 0.17 | 0.91 | 0.78 | 0.33 | 0.11 | 0.99 | 0.81 |
| **Spatial Nback 2-1back** | 0.13 | -0.44 | 1.11 | 0.90 | 0.32 | 0.08 | 0.94 | 0.76 | 0.30 | 0.04 | 1.02 | 0.82 |
| **Spatial Nback 1back** | 0.50 | -0.39 | 1.09 | 0.87 | 0.20 | 0.01 | 0.97 | 0.83 | 0.28 | 0.03 | 1.03 | 0.85 |
| **Multitask FC** | -0.26 | -0.62 | 1.19 | 0.92 | 0.51 | 0.16 | 0.92 | 0.79 | 0.23 | 0.05 | 1.02 | 0.84 |
| **Subcortical volume** | 0.19 | -0.27 | 0.97 | 0.83 | 0.18 | -0.07 | 0.99 | 0.83 | 0.23 | 0.05 | 0.98 | 0.83 |
| **FreeSurfer summations** | 0.30 | -0.25 | 0.96 | 0.82 | 0.09 | -0.11 | 1.01 | 0.86 | 0.20 | 0.04 | 0.99 | 0.84 |
| **Verbal Nback 2-1back** | 0.05 | -0.60 | 1.08 | 0.89 | 0.21 | 0.03 | 0.94 | 0.81 | 0.19 | 0.00 | 1.00 | 0.84 |
| **Cortical thickness** | 0.09 | -0.32 | 0.99 | 0.79 | 0.12 | -0.10 | 1.00 | 0.85 | 0.17 | 0.02 | 1.00 | 0.82 |
| **Spatial Nback FC** | -0.03 | -0.57 | 1.16 | 0.95 | 0.29 | 0.06 | 0.97 | 0.83 | 0.16 | -0.01 | 1.04 | 0.87 |
| **Verbal Nback 1back** | -0.01 | -0.65 | 1.10 | 0.86 | 0.08 | -0.03 | 0.97 | 0.83 | 0.09 | -0.05 | 1.03 | 0.84 |

|  | **ADHD** | | | | **non-ADHD** | | | | **All** | | | |
| --- | --- | --- | --- | --- | --- | --- | --- | --- | --- | --- | --- | --- |
|  | **r** | **R2** | **MAE** | **RMSE** | **r** | **R2** | **MAE** | **RMSE** | **r** | **R2** | **MAE** | **RMSE** |
| **Spatial Nback Contrasts** | 0.42 | -0.33 | 1.07 | 0.87 | 0.36 | 0.12 | 0.92 | 0.76 | 0.44 | 0.10 | 0.99 | 0.81 |
| **sMRI + Verbal Nback FC + Contrasts** | 0.25 | -0.25 | 0.96 | 0.77 | 0.33 | 0.07 | 0.92 | 0.78 | 0.38 | 0.13 | 0.94 | 0.77 |
| **sMRI + G-Multitask FC + Spatial Nback Contrasts** | 0.36 | -0.19 | 0.94 | 0.76 | 0.33 | 0.03 | 0.94 | 0.78 | 0.37 | 0.12 | 0.94 | 0.77 |
| **sMRI + G-Multitask FC + Verbal Nback Contrasts** | 0.12 | -0.34 | 1.00 | 0.82 | 0.38 | 0.09 | 0.91 | 0.78 | 0.36 | 0.11 | 0.95 | 0.80 |
| **sMRI + Spatial Nback FC + Contrasts** | 0.38 | -0.19 | 0.94 | 0.73 | 0.23 | -0.01 | 0.96 | 0.79 | 0.34 | 0.10 | 0.95 | 0.76 |
| **sMRI + Multitask FC + Verbal Nback Contrasts** | 0.12 | -0.54 | 1.07 | 0.85 | 0.38 | 0.14 | 0.89 | 0.75 | 0.34 | 0.07 | 0.97 | 0.80 |
| **sMRI + Multitask FC + Spatial Nback Contrasts** | 0.26 | -0.39 | 1.01 | 0.80 | 0.30 | 0.04 | 0.94 | 0.77 | 0.31 | 0.07 | 0.97 | 0.78 |
| **Verbal Nback Contrasts** | 0.06 | -0.68 | 1.11 | 0.88 | 0.31 | 0.09 | 0.91 | 0.77 | 0.28 | 0.01 | 1.00 | 0.81 |
| **sMRI** | 0.17 | -0.32 | 0.99 | 0.78 | 0.18 | -0.14 | 1.02 | 0.84 | 0.20 | 0.00 | 1.01 | 0.81 |

**Supplementary Table 6.** Performance metrics of second layer stacked modalities across non-adhd and adhd tiers in external validation

| **Network Pair** | **Average Coefficient** | **std** |
| --- | --- | --- |
| **Dorsal_Attention-Posterior_Multimodal** | 0.0009 | 0.0005 |
| **Cingulo-Opercular-Posterior_Multimodal** | 0.0008 | 0.0005 |
| **Visual1-Visual2** | 0.0008 | 0.0005 |
| **Ventral_Multimodal-Visual2** | 0.0008 | 0.0003 |
| **Posterior_Multimodal-Visual2** | 0.0008 | 0.0004 |
| **Dorsal_Attention-Visual2** | 0.0007 | 0.0004 |
| **Subcortex-Visual2** | 0.0007 | 0.0004 |
| **Default-Frontoparietal** | 0.0007 | 0.0008 |
| **Cingulo-Opercular-Visual2** | 0.0007 | 0.0005 |
| **Dorsal_Attention-Ventral_Multimodal** | 0.0006 | 0.0003 |
| **Ventral_Multimodal-Visual1** | 0.0006 | 0.0002 |
| **Language-Posterior_Multimodal** | 0.0006 | 0.0005 |
| **Language-Ventral_Multimodal** | 0.0006 | 0.0003 |
| **Posterior_Multimodal-Ventral_Multimodal** | 0.0006 | 0.0001 |
| **Dorsal_Attention-Subcortex** | 0.0006 | 0.0005 |
| **Cingulo-Opercular-Subcortex** | 0.0006 | 0.0005 |
| **Cingulo-Opercular-Somatomotor** | 0.0005 | 0.0007 |
| **Cingulo-Opercular-Ventral_Multimodal** | 0.0005 | 0.0003 |
| **Posterior_Multimodal-Subcortex** | 0.0005 | 0.0003 |
| **Somatomotor-Subcortex** | 0.0005 | 0.0005 |
| **Dorsal_Attention-Language** | 0.0005 | 0.0005 |
| **Language-Visual2** | 0.0004 | 0.0006 |
| **Cingulo-Opercular-Dorsal_Attention** | 0.0004 | 0.0006 |
| **Dorsal_Attention-Visual1** | 0.0004 | 0.0006 |
| **Default-Dorsal_Attention** | 0.0004 | 0.0008 |
| **Posterior_Multimodal-Visual1** | 0.0004 | 0.0004 |
| **Default-Language** | 0.0004 | 0.0007 |
| **Cingulo-Opercular-Visual1** | 0.0004 | 0.0005 |
| **Orbito-Affective-Visual2** | 0.0003 | 0.0005 |
| **Language-Subcortex** | 0.0003 | 0.0005 |
| **Cingulo-Opercular-Language** | 0.0003 | 0.0007 |
| **Subcortex-Visual1** | 0.0003 | 0.0004 |
| **Somatomotor-Visual2** | 0.0003 | 0.0004 |
| **Dorsal_Attention-Orbito-Affective** | 0.0002 | 0.0004 |
| **Orbito-Affective-Posterior_Multimodal** | 0.0002 | 0.0004 |
| **Default-Visual2** | 0.0002 | 0.0006 |
| **Dorsal_Attention-Frontoparietal** | 0.0002 | 0.0007 |
| **Frontoparietal-Language** | 0.0002 | 0.0006 |
| **Cingulo-Opercular-Default** | 0.0002 | 0.0007 |
| **Cingulo-Opercular-Orbito-Affective** | 0.0002 | 0.0005 |
| **Somatomotor-Ventral_Multimodal** | 0.0002 | 0.0003 |
| **Language-Visual1** | 0.0002 | 0.0006 |
| **Frontoparietal-Posterior_Multimodal** | 0.0001 | 0.0007 |
| **Default-Subcortex** | 0.0001 | 0.0006 |
| **Orbito-Affective-Ventral_Multimodal** | 0.0001 | 0.0004 |
| **Subcortex-Ventral_Multimodal** | 0.0001 | 0.0004 |
| **Language-Orbito-Affective** | 0.0001 | 0.0004 |
| **Frontoparietal-Subcortex** | 0.0001 | 0.0005 |
| **Auditory-Somatomotor** | 0.0000 | 0.0005 |
| **Orbito-Affective-Somatomotor** | 0.0000 | 0.0005 |
| **Default-Orbito-Affective** | 0.0000 | 0.0005 |
| **Auditory-Cingulo-Opercular** | 0.0000 | 0.0005 |
| **Orbito-Affective-Visual1** | 0.0000 | 0.0004 |
| **Default-Ventral_Multimodal** | 0.0000 | 0.0006 |
| **Frontoparietal-Ventral_Multimodal** | -0.0001 | 0.0005 |
| **Somatomotor-Visual1** | -0.0001 | 0.0004 |
| **Posterior_Multimodal-Somatomotor** | -0.0001 | 0.0005 |
| **Default-Posterior_Multimodal** | -0.0001 | 0.0005 |
| **Orbito-Affective-Subcortex** | -0.0001 | 0.0006 |
| **Frontoparietal-Orbito-Affective** | -0.0001 | 0.0004 |
| **Dorsal_Attention-Somatomotor** | -0.0002 | 0.0005 |
| **Auditory-Subcortex** | -0.0002 | 0.0003 |
| **Default-Visual1** | -0.0002 | 0.0006 |
| **Frontoparietal-Visual2** | -0.0002 | 0.0007 |
| **Auditory-Visual2** | -0.0003 | 0.0004 |
| **Cingulo-Opercular-Frontoparietal** | -0.0003 | 0.0008 |
| **Language-Somatomotor** | -0.0003 | 0.0006 |
| **Auditory-Ventral_Multimodal** | -0.0004 | 0.0003 |
| **Default-Somatomotor** | -0.0004 | 0.0005 |
| **Auditory-Orbito-Affective** | -0.0004 | 0.0004 |
| **Auditory-Visual1** | -0.0005 | 0.0004 |
| **Frontoparietal-Visual1** | -0.0005 | 0.0006 |
| **Auditory-Posterior_Multimodal** | -0.0005 | 0.0003 |
| **Auditory-Dorsal_Attention** | -0.0005 | 0.0005 |
| **Auditory-Default** | -0.0006 | 0.0005 |
| **Auditory-Language** | -0.0007 | 0.0005 |
| **Frontoparietal-Somatomotor** | -0.0008 | 0.0006 |
| **Auditory-Frontoparietal** | -0.0009 | 0.0005 |

**Supplementary Table 7.** The average elastic net coefficients (across test sets) for connectivity between regions in each pair of networks for mid fc.

| **Region** | **Weight** | **Region** | **Weight** | **Region** | **Weight** | **Region** | **Weight** |
| --- | --- | --- | --- | --- | --- | --- | --- |
| R_PCV | 0.023 | R_d32 | 0.007 | putamen_left | 0.005 | L_TGd | 0.002 |
| L_PCV | 0.018 | R_7PL | 0.007 | R_PGs | 0.005 | R_ProS | 0.002 |
| R_a32pr | 0.018 | R_IP1 | 0.007 | L_STSdp | 0.005 | L_FFC | 0.002 |
| R_AVI | 0.018 | L_7PL | 0.007 | R_VVC | 0.004 | R_MBelt | 0.002 |
| L_6a | 0.017 | R_8BM | 0.007 | R_a10p | 0.004 | R_6ma | 0.002 |
| R_7Pm | 0.015 | R_p24 | 0.007 | R_1 | 0.004 | L_V2 | 0.002 |
| R_i6-8 | 0.015 | R_IP2 | 0.007 | L_IFJa | 0.004 | caudate_left | 0.002 |
| R_7Am | 0.014 | L_PSL | 0.007 | R_TE1m | 0.004 | R_FOP4 | 0.002 |
| R_6a | 0.014 | R_V6 | 0.006 | L_6d | 0.004 | L_31pd | 0.002 |
| L_a32pr | 0.013 | L_IFSa | 0.006 | L_IFSp | 0.004 | R_47m | 0.002 |
| L_10v | 0.012 | L_i6-8 | 0.006 | L_IP0 | 0.004 | L_V8 | 0.002 |
| L_AIP | 0.012 | L_p32pr | 0.006 | R_PGp | 0.004 | L_8Ad | 0.002 |
| L_AVI | 0.012 | R_FEF | 0.006 | brainStem | 0.004 | L_PEF | 0.002 |
| L_FEF | 0.012 | L_33pr | 0.006 | amygdala_right | 0.004 | R_STV | 0.002 |
| R_33pr | 0.012 | R_H | 0.006 | L_TF | 0.004 | R_5L | 0.002 |
| L_IP2 | 0.011 | L_d32 | 0.006 | R_PreS | 0.004 | L_TA2 | 0.002 |
| L_SCEF | 0.011 | L_13l | 0.006 | R_PGi | 0.004 | R_V1 | 0.002 |
| R_46 | 0.011 | R_p24pr | 0.006 | R_PoI2 | 0.004 | L_PHA1 | 0.002 |
| L_PreS | 0.010 | putamen_right | 0.006 | L_TPOJ2 | 0.004 | R_A1 | 0.002 |
| L_55b | 0.010 | L_STV | 0.005 | L_44 | 0.004 | R_44 | 0.002 |
| L_7m | 0.010 | thalamus_right | 0.005 | thalamus_left | 0.004 | L_A1 | 0.002 |
| L_s6-8 | 0.010 | R_7m | 0.005 | L_LO2 | 0.004 | L_MT | 0.002 |
| L_EC | 0.009 | L_FOP5 | 0.005 | L_VVC | 0.004 | L_TPOJ3 | 0.002 |
| L_PeEc | 0.009 | R_TA2 | 0.005 | R_p32pr | 0.004 | R_A4 | 0.002 |
| L_PGi | 0.009 | R_Ig | 0.005 | R_6mp | 0.004 | L_25 | 0.002 |
| cerebellum_right | 0.008 | L_FOP4 | 0.005 | L_RSC | 0.003 | pallidum_right | 0.001 |
| L_6r | 0.008 | L_a9-46v | 0.005 | R_pOFC | 0.003 | R_PEF | 0.001 |
| L_7Am | 0.008 | L_V3B | 0.005 | R_p32 | 0.003 | L_VMV1 | 0.002 |
| L_7Pm | 0.008 | R_PHT | 0.005 | R_p9-46v | 0.003 | R_OFC | 0.002 |
| L_MIP | 0.008 | R_VMV1 | 0.005 | diencephalon_left | 0.003 | R_8Av | 0.001 |
| L_ProS | 0.008 | L_V4t | 0.005 | R_5mv | 0.003 | R_PeEc | 0.001 |
| L_SFL | 0.008 | R_RSC | 0.005 | L_a24pr | 0.003 | hippocampus_right | 0.001 |
| R_55b | 0.008 | L_p24 | 0.005 | L_8BM | 0.003 | L_6ma | 0.001 |
| R_A5 | 0.008 | L_IFJp | 0.005 | L_a10p | 0.003 | R_VMV2 | 0.001 |
| R_AIP | 0.008 | R_25 | 0.005 | L_STSva | 0.003 | R_IP0 | 0.001 |
| R_p10p | 0.008 | L_p10p | 0.005 | R_OP2-3 | 0.003 | R_POS2 | 0.002 |
| R_Pir | 0.008 | L_p9-46v | 0.005 | L_11l | 0.003 | R_6v | 0.001 |
| R_VIP | 0.008 | cerebellum_left | 0.005 | R_MIP | 0.003 | L_OP4 | 0.001 |
| L_46 | 0.007 | L_FST | 0.005 | R_s6-8 | 0.003 | amygdala_left | 0.001 |
| R_EC | 0.007 | R_FOP5 | 0.005 | L_LIPd | 0.003 | R_a47r | 0.001 |
| R_PFm | 0.007 | R_LIPv | 0.005 | R_MT | 0.002 | L_LBelt | 0.001 |
| L_47m | 0.001 | L_2 | -0.001 | R_13l | -0.003 | R_TPOJ2 | -0.004 |
| L_V6 | 0.001 | L_s32 | -0.001 | R_TE1p | -0.003 | R_47s | -0.004 |
| pallidum_left | 0.001 | R_TGd | -0.001 | L_V3CD | -0.003 | R_PH | -0.004 |
| R_7PC | 0.001 | R_43 | -0.001 | R_DVT | -0.003 | R_TE2a | -0.004 |
| R_LO3 | 0.001 | R_TPOJ3 | -0.001 | R_STSdp | -0.003 | R_24dv | -0.005 |
| diencephalon_right | 0.000 | L_PHT | -0.001 | L_PHA3 | -0.003 | L_a47r | -0.005 |
| L_9-46d | 0.000 | L_VMV3 | -0.001 | L_TE1m | -0.003 | R_PF | -0.005 |
| L_LO3 | 0.000 | L_9m | -0.001 | R_6r | -0.003 | R_9-46d | -0.005 |
| L_MI | 0.000 | L_LIPv | -0.001 | R_V3 | -0.003 | L_TPOJ1 | -0.005 |
| L_OP2-3 | 0.000 | R_V8 | -0.001 | R_STSva | -0.003 | R_MI | -0.004 |
| L_PIT | 0.000 | L_PFt | -0.001 | R_LBelt | -0.003 | R_V2 | -0.005 |
| L_PoI2 | 0.000 | L_4 | -0.001 | L_a24 | -0.003 | R_OP4 | -0.005 |
| L_STGa | 0.000 | L_V1 | -0.001 | L_IPS1 | -0.003 | L_V6A | -0.005 |
| L_TE1a | 0.000 | L_RI | -0.001 | L_p24pr | -0.003 | R_STSda | -0.005 |
| R_10v | 0.000 | R_LO2 | -0.001 | R_d23ab | -0.003 | R_9p | -0.005 |
| R_23d | 0.000 | accumbens_right | -0.001 | R_IFJp | -0.003 | R_LIPd | -0.005 |
| R_31a | 0.000 | L_V4 | -0.001 | L_LO1 | -0.003 | R_V3CD | -0.005 |
| R_6d | 0.000 | R_FST | -0.001 | R_10pp | -0.003 | L_PBelt | -0.005 |
| R_FFC | 0.000 | L_V7 | -0.001 | L_IP1 | -0.003 | R_31pd | -0.005 |
| R_IFJa | 0.000 | L_5L | -0.001 | R_V4t | -0.003 | L_8BL | -0.005 |
| R_PI | 0.000 | R_8Ad | -0.001 | R_OP1 | -0.003 | L_TGv | -0.005 |
| R_RI | 0.000 | R_LO1 | -0.001 | L_VMV2 | -0.003 | R_2 | -0.005 |
| R_SCEF | 0.000 | caudate_right | -0.001 | R_10r | -0.003 | L_DVT | -0.006 |
| L_1 | 0.000 | R_23c | -0.002 | R_IPS1 | -0.004 | L_FOP2 | -0.005 |
| L_23c | 0.000 | L_5mv | -0.002 | L_3b | -0.003 | L_p47r | -0.005 |
| L_3a | 0.000 | R_7AL | -0.002 | R_5m | -0.003 | L_STSda | -0.006 |
| L_9p | 0.000 | R_PFcm | -0.002 | L_TE1p | -0.004 | L_FOP1 | -0.005 |
| L_TE2a | 0.000 | L_OP1 | -0.002 | L_FOP3 | -0.003 | L_10d | -0.005 |
| L_VIP | 0.000 | L_V3 | -0.002 | L_p32 | -0.004 | R_10d | -0.005 |
| R_4 | 0.000 | L_23d | -0.002 | hippocampus_left | -0.003 | R_FOP3 | -0.005 |
| R_PIT | 0.000 | L_45 | -0.002 | L_MST | -0.004 | R_PFt | -0.005 |
| R_STGa | 0.000 | L_POS2 | -0.002 | L_7AL | -0.003 | L_8C | -0.005 |
| R_TF | 0.000 | L_PFm | -0.002 | R_a24pr | -0.003 | L_PGs | -0.006 |
| accumbens_left | -0.001 | R_TE2p | -0.002 | R_PBelt | -0.004 | R_TPOJ1 | -0.006 |
| L_6mp | -0.001 | R_PHA2 | -0.002 | R_V4 | -0.004 | L_Pir | -0.006 |
| L_6v | -0.001 | R_3a | -0.002 | L_A4 | -0.004 | R_VMV3 | -0.006 |
| L_7PC | -0.001 | L_47s | -0.002 | L_pOFC | -0.004 | L_H | -0.006 |
| L_Ig | -0.001 | R_11l | -0.002 | L_10r | -0.004 | R_STSvp | -0.006 |
| L_PGp | -0.001 | R_52 | -0.002 | R_TE1a | -0.004 | R_AAIC | -0.006 |
| L_TE2p | -0.001 | R_TGv | -0.002 | L_43 | -0.004 | R_p47r | -0.006 |
| R_3b | -0.001 | R_FOP2 | -0.002 | R_V3B | -0.004 | R_a9-46v | -0.006 |
| R_PHA1 | -0.001 | L_V3A | -0.002 | R_IFSp | -0.004 | L_PFcm | -0.006 |
| L_PH | -0.001 | R_SFL | -0.015 | L_PF | -0.004 | L_10pp | -0.007 |
| L_A5 | -0.002 | L_PoI1 | -0.016 | R_s32 | -0.004 | L_31a | -0.007 |
| L_5m | -0.007 |  |  |  |  |  |  |
| L_9a | -0.007 |  |  |  |  |  |  |
| L_STSvp | -0.007 |  |  |  |  |  |  |
| R_MST | -0.007 |  |  |  |  |  |  |
| R_POS1 | -0.007 |  |  |  |  |  |  |
| R_PSL | -0.007 |  |  |  |  |  |  |
| R_V3A | -0.007 |  |  |  |  |  |  |
| R_V7 | -0.007 |  |  |  |  |  |  |
| L_24dd | -0.008 |  |  |  |  |  |  |
| L_52 | -0.008 |  |  |  |  |  |  |
| L_AAIC | -0.008 |  |  |  |  |  |  |
| L_OFC | -0.008 |  |  |  |  |  |  |
| L_PHA2 | -0.008 |  |  |  |  |  |  |
| R_a24 | -0.008 |  |  |  |  |  |  |
| R_V6A | -0.008 |  |  |  |  |  |  |
| L_MBelt | -0.009 |  |  |  |  |  |  |
| L_PFop | -0.009 |  |  |  |  |  |  |
| L_POS1 | -0.009 |  |  |  |  |  |  |
| R_24dd | -0.009 |  |  |  |  |  |  |
| R_9a | -0.009 |  |  |  |  |  |  |
| R_FOP1 | -0.009 |  |  |  |  |  |  |
| R_IFSa | -0.009 |  |  |  |  |  |  |
| L_8Av | -0.010 |  |  |  |  |  |  |
| R_45 | -0.010 |  |  |  |  |  |  |
| R_9m | -0.010 |  |  |  |  |  |  |
| R_PFop | -0.010 |  |  |  |  |  |  |
| R_v23ab | -0.010 |  |  |  |  |  |  |
| L_31pv | -0.011 |  |  |  |  |  |  |
| R_8C | -0.011 |  |  |  |  |  |  |
| L_47l | -0.012 |  |  |  |  |  |  |
| L_PI | -0.012 |  |  |  |  |  |  |
| R_31pv | -0.012 |  |  |  |  |  |  |
| L_d23ab | -0.013 |  |  |  |  |  |  |
| R_47l | -0.013 |  |  |  |  |  |  |
| R_PoI1 | -0.013 |  |  |  |  |  |  |
| L_24dv | -0.014 |  |  |  |  |  |  |
| R_PHA3 | -0.016 |  |  |  |  |  |  |
| R_8BL | -0.017 |  |  |  |  |  |  |
| L_v23ab | -0.019 |  |  |  |  |  |  |

**Supplementary Table 8.** Average Elastic Net coefficients (across test sets) based on Glasser parcellations for 2-0back contrast.

| **Region** | **Weight** | **Region** | **Weight** | **Region** | **Weight** |
| --- | --- | --- | --- | --- | --- |
| b'G_and_S_paracentral' | 0.030 | b'G_parietal_sup' | 0.007 | b'G_insular_short' | -0.003 |
| b'S_cingul-Marginalis' | 0.026 | b'S_pericallosal' | 0.007 | b'G_and_S_cingul-Mid-Post' | -0.003 |
| b'S_orbital_med-olfact' | 0.025 | b'S_oc-temp_lat' | 0.007 | b'G_oc-temp_med-Lingual' | -0.003 |
| b'S_calcarine' | 0.022 | b'G_pariet_inf-Supramar' | 0.007 | b'Lat_Fis-ant-Horizont' | -0.012 |
| b'G_and_S_transv_frontopol' | 0.021 | b'Pole_temporal' | 0.006 | b'G_and_S_transv_frontopol' | -0.012 |
| b'G_and_S_cingul-Mid-Ant' | 0.018 | b'S_suborbital' | 0.004 | b'G_cuneus' | -0.012 |
| b'S_front_inf' | 0.015 | b'G_pariet_inf-Supramar' | 0.004 | b'G_orbital' | -0.013 |
| b'G_temporal_inf' | 0.015 | b'Lat_Fis-ant-Vertical' | 0.004 | b'Lat_Fis-post' | -0.014 |
| b'G_oc-temp_med-Lingual' | 0.015 | b'S_temporal_transverse' | 0.003 | b'G_and_S_cingul-Mid-Post' | -0.014 |
| b'S_occipital_ant' | 0.015 | b'G_insular_short' | 0.003 | b'G_oc-temp_lat-fusifor' | -0.015 |
| b'G_and_S_frontomargin' | 0.014 | b'G_front_inf-Triangul' | 0.003 | b'G_cingul-Post-dorsal' | -0.015 |
| b'S_cingul-Marginalis' | 0.014 | b'G_and_S_frontomargin' | 0.003 | b'G_temp_sup-G_T_transv' | -0.015 |
| b'S_circular_insula_sup' | 0.013 | b'S_temporal_sup' | 0.003 | b'G_front_inf-Opercular' | -0.016 |
| b'S_temporal_inf' | 0.013 | b'Pole_temporal' | 0.002 | b'S_central' | -0.017 |
| b'G_postcentral' | 0.013 | b'Pole_occipital' | 0.002 | b'S_front_sup' | -0.017 |
| b'G_front_middle' | 0.013 | b'S_postcentral' | 0.002 | b'S_oc_sup_and_transversal' | -0.017 |
| b'S_central' | 0.013 | b'G_precentral' | 0.002 | b'G_temp_sup-Lateral' | -0.017 |
| b'S_collat_transv_ant' | 0.012 | b'S_precentral-inf-part' | 0.001 | b'G_front_inf-Triangul' | -0.018 |
| b'S_temporal_sup' | 0.012 | b'G_occipital_middle' | 0.001 | b'S_front_middle' | -0.018 |
| b'S_circular_insula_ant' | 0.012 | b'S_orbital-H_Shaped' | 0.001 | b'Lat_Fis-ant-Horizont' | -0.019 |
| b'G_orbital' | 0.012 | b'S_orbital_lateral' | 0.001 | b'G_oc-temp_med-Parahip' | -0.020 |
| b'S_interm_prim-Jensen' | 0.011 | b'Lat_Fis-post' | 0.000 | b'S_oc-temp_lat' | -0.021 |
| b'S_subparietal' | 0.011 | b'G_front_inf-Orbital' | 0.000 | b'G_cingul-Post-ventral' | -0.021 |
| b'G_parietal_sup' | 0.011 | b'G_occipital_middle' | 0.000 | b'S_circular_insula_ant' | -0.021 |
| b'G_oc-temp_lat-fusifor' | 0.011 | b'Medial_wall' | 0.000 | b'G_cingul-Post-dorsal' | -0.022 |
| b'S_oc_middle_and_Lunatus' | 0.011 | b'Medial_wall' | 0.000 | b'G_and_S_occipital_inf' | -0.022 |
| b'G_rectus' | 0.011 | b'G_subcallosal' | 0.000 | b'S_intrapariet_and_P_trans' | -0.022 |
| b'S_occipital_ant' | 0.010 | b'S_temporal_inf' | -0.001 | b'G_and_S_cingul-Ant' | -0.022 |
| b'G_temp_sup-Lateral' | 0.010 | b'G_front_sup' | -0.001 | b'S_precentral-sup-part' | -0.025 |
| b'G_temp_sup-Plan_polar' | 0.010 | b'G_precuneus' | -0.001 | b'S_collat_transv_post' | -0.027 |
| b'S_parieto_occipital' | 0.008 | b'G_Ins_lg_and_S_cent_ins' | -0.001 | b'G_temp_sup-Plan_polar' | -0.027 |
| b'G_and_S_cingul-Ant' | 0.008 | b'G_front_inf-Orbital' | -0.001 | b'S_oc_sup_and_transversal' | -0.031 |
| b'G_temporal_middle' | 0.008 | b'G_postcentral' | -0.001 | b'G_temp_sup-G_T_transv' | -0.033 |
| b'S_pericallosal' | 0.008 | b'S_front_middle' | -0.001 | b'G_and_S_subcentral' | -0.036 |
| b'Pole_occipital' | 0.008 | b'G_cuneus' | -0.001 |  |  |
| b'S_oc-temp_med_and_Lingual' | 0.008 | b'G_front_sup' | -0.002 |  |  |
| b'G_rectus' | 0.008 | b'G_precuneus' | -0.002 |  |  |
| b'G_pariet_inf-Angular' | 0.008 | b'G_Ins_lg_and_S_cent_ins' | -0.002 |  |  |
| b'G_subcallosal' | 0.008 | b'G_temporal_middle' | -0.002 |  |  |
| b'S_suborbital' | 0.008 | b'G_front_middle' | -0.002 |  |  |

**Supplementary Table 9.** Average Elastic Net coefficients (across test sets) based on Gordon parcellations for T2 Gray Matter Intensity.

| First level categories of the ATC medication system | Number of Participants |
| --- | --- |
| Alimentary tract and metabolism | 144 |
| Blood and blood forming organs | 12 |
| Cardiovascular system | 124 |
| Dermatologicals | 108 |
| Genitourinary system and sex hormones | 72 |
| Systemic hormonal preparations, excl. sex hormones and insulins | 26 |
| Anti-infectives for systemic use | 56 |
| Antineoplastic and immunomodulating agents | 5 |
| Musculo-skeletal system | 145 |
| Nervous system | 710 |
| Antiparasitic products, insecticides and repellents | 5 |
| Respiratory system | 721 |
| Sensory organs | 42 |
| Various | 1 |

**Supplementary Table 10.** The number of participants in ABCD study with exposure to each of the 14 first-level categories of the Anatomical Therapeutic Chemical (ATC) system.
