## Supplementary Methods and Supplementary Figures for "Predicting Cognitive Functioning in ADHD Using Population-Based MRI Across Large and Small Samples"

#### Attention-Deficit/Hyperactivity Disorder (ADHD) Identification

To filter participants based on ADHD symptoms, we applied the tiered selection method developed by Cordova et al. (Cordova et al., 2022). This method approximates The Diagnostic and Statistical Manual of Mental Illnesses (DSM-5) (American Psychiatric Association, 2022) criteria for present ADHD symptoms using available mental health measures from the Adolescent Brain Cognitive Development (ABCD) dataset and defines four increasingly stringent tiers (Tiers 1–4) of ADHD criteria including parent and teacher reports and rule out information for comorbid conditions. While Cordova and colleagues used multiple imputation to retain the original sample size, we opted to analyze the available data without imputation due to approximately 70% missing data on teacher-rated Brief Problem Monitoring (BPM) ratings. Groups were formed using measures from the Computerized Schedule of Affective Disorders and Schizophrenia for School-Age Children (K-SADS-COMP) (Townsend et al., 2020), parent-reported Child Behavior Checklist (CBCL) (Achenbach & Ruffle, 2000), teacher-reported Brief Problem Monitor (BPM) (Achenbach & Rescorla, 2001), and rule-out information:

1) Tier 1 included participants who met the criteria for only current ADHD according to the parent-reported K-SADS-COMP.
2) Tier 2 excluded Tier 1 participants who had a diagnosis of bipolar disorder, unspecified schizophrenia spectrum (per K-SADS-COMP), and/or intellectual disability based on NIH Toolbox WISC-V Matrix Reasoning (Wechsler, 2014).
3) Tier 3 used the teacher-BPM attention problems scale to confirm that ADHD symptoms were evident in more than one setting.
4) Tier 4 added the clinical cutoff from the parent CBCL attention scale or the CBCL ADHD DSM-5 scale as an additional diagnostic criterion.

### Cognitive-Functioning

For the ABCD, NIH Toolbox composite scores (Akshoomoff et al., 2013) for total cognitive-functioning were derived from seven tasks: Picture Vocabulary measures receptive vocabulary and verbal comprehension (Gershon et al., 2013). Flanker Inhibitory Control & Attention evaluates attention and response inhibition (Zelazo et al., 2013). List Sorting Working Memory tests working memory and sequencing (Tulsky et al., 2013). Dimensional Change Card Sort assesses cognitive flexibility and set-shifting (Zelazo et al., 2013). Pattern Comparison Processing Speed measures visual processing speed (Carlozzi et al., 2013). Picture Sequence Memory evaluates episodic memory and sequential recall (Bauer et al., 2013). Oral Reading Recognition tests reading ability and word recognition (Gershon et al., 2013). Together, these tasks provide a comprehensive assessment of cognitive function across multiple domains.

For Lytle and colleagues’, The Wechsler Abbreviated Scale of Intelligence, Second Edition (WASI-II) (Wechsler, 1999) was used to measure cognitive functioning. It includes four subtests that provide a composite full-scale IQ score. Vocabulary assesses word knowledge and verbal comprehension. Similarities measures abstract reasoning and conceptual thinking. Block Design evaluates spatial visualization and problem-solving. Matrix Reasoning tests nonverbal reasoning and pattern recognition. Together, these subtests offer a broad assessment of cognitive ability (McCrimmon & Smith, 2013).

### Neuroimaging: ABCD

### Quality Considerations

We excluded Site 22 because it only had data from 32 children (Garavan et al., 2018). In accordance with the release notes (Yang & Jernigan), we corrected site IDs for 25 children due to entry errors. We also excluded 58 children who had vision problems (Luciana et al., 2018).

Quality control for neuroimaging data followed ABCD-recommended exclusion criteria (Jernigan et al., 2023). Exclusion flags based on metrics such as image quality, MR neurological screenings, and task performance, were assigned to each neuroimaging feature (Hagler et al., 2019). We did not apply listwise deletion, but rather excluded specific sets of neuroimaging features that failed to meet the criteria. For instance, when participants failed to meet the criteria for Nback task fMRI, but not other sets, we only dropped Nback task fMRI from our analysis. See below under Cognitive Prediction Models on how we dealt with missingness through opportunistic stacking (Pat et al., 2022).

ABCD provided post-processed tabulated data with specific atlases. And we use them directly. We then also post-processed the ABCD-BIDS Community Collection (ABCC) data ourselves to complement the post-processed data provided by the ABCD.

#### ABCC Pre-processing Pipeline

ABCC is a curated dataset derived from the Adolescent Brain Cognitive Development (ABCD) Study, formatted according to the Brain Imaging Data Structure (BIDS) standard (Scott et al., 2020). It provides easy-to-use neuroimaging and behavioral data for researchers. The ABCC includes preprocessed data like MRI images and associated metadata. This collection also includes the outputs of fMRIPrep (version: 20.2.0) and the Human Connectome Project's minimal preprocessing pipelines. We used the generated confounds table from fMRIPrep and the CIFTI file from hcp-pipeline outputs. We converted the text E-prime event files to tsv files using the eprimetotsv.py script by Demidenko and colleagues (Demidenko, 2024). The resulting event times are calculated from the task start time after calibration scans. Our first level analysis pipeline was mainly built with Nilearn (Abraham et al., 2014), Nibabel (Brett et al., 2024) packages .

For the task fMRI in the ABCC, we conducted the following processing steps. First, nuisance regressors were selected and included in the design matrix based on recommendations in the literature (Ciric et al., 2017; Gotts et al., 2013). These include a subset of confounds from fMRIPrep to account for potential noise sources: Cosine regressors were added to remove low-frequency noise, such as scanner drift or physiological cycles, which are unrelated to neural activity (Ciric et al., 2017). Additionally, ten anatomical CompCor components were derived from signals within anatomically defined noise regions, including white matter and cerebrospinal fluid (CSF). These principal components (PC1–PC10), which are unlikely to contain neural activity, were included to account for physiological noise like heartbeat and respiration (Muschelli et al., 2014). Movement regressors, representing head motion parameters calculated during realignment, were also included. These regressors consisted of translations (x, y, z) and rotations (roll, pitch, yaw) in three dimensions, as well as their first derivatives (rate of change). Including these regressors mitigates motion-related artifacts, such as spurious correlations and signal distortions caused by subject movement during the scan (Maknojia et al., 2019). Specific regressors included rot_x, rot_x_derivative1, rot_y, rot_y_derivative1, rot_z, rot_z_derivative1, trans_x, trans_x_derivative1, trans_y, trans_y_derivative1, trans_z, and trans_z_derivative1. To address initial calibration effects, dummy scans were removed from the confounds and image data. The number of removed TRs varied by scanner type: Siemens and Philips scans had 8 TRs removed, GE DV25 scans had 5, and GE DV26 scans had 16 TRs removed (Jernigan et al., 2023). Frame times were calculated based on the number of scans and TR.

The first-level design matrix was created using the SPM HRF model. Subsequently, time series standardization was performed, and a General Linear Model (GLM) analysis was run using Ordinary Least Squares (OLS) as the noise model. Contrasts of interest were defined by creating a contrast matrix. For each contrast, effect sizes were computed by estimating the GLM coefficients. These results were averaged by computing fixed effects, incorporating variance across runs, and calculating the weighted average effect size for contrasts across the two runs. Finally, the averaged effect size maps were parcellated into 379 regions, comprising 360 cortical regions based on the Glasser atlas (Glasser et al., 2016) and 19 FreeSurfer (Fischl et al., 2002) subcortical regions.

For the task and rest fMRI FC in the ABCC, we conducted the following processing steps. We began with the selection of nuisance regressors and the removal of dummy scans, as described earlier. Frame times were calculated based on the number of scans and TRs. Outlier detection and regression were applied by identifying high-motion TRs with framewise displacement (FD) > 0.5 and standardized derivative of root mean square variance over voxels (DVARS) > 1.5, as described in prior literature (Phạm et al., 2023; Power et al., 2013). FD was calculated as the sum of the absolute displacements in all six motion parameters (translation along x, y, z axes, and rotation about these axes) between successive fMRI volumes. DVARS quantified the root mean square of signal intensity differences across the brain between successive volumes (Afyouni & Nichols, 2018). The identified TRs were flagged as outliers and added as additional nuisance regressors (Power et al., 2014). A censoring mask was created to exclude flagged outliers, their adjacent five TRs, and any time intervals containing fewer than five consecutive sub-threshold TRs. Participants were excluded if their censoring mask flagged more than half of the total timepoints. A lenient FD threshold was used, with no additional motion scrubbing, due to the multiband acquisition, participant demographics, and the importance of preserving information when examining brain-behavior associations (Addeh et al., 2023; Phạm et al., 2023; Power et al., 2019).

The first-level design matrix was constructed using the SPM HRF model for task fMRI, while no HRF model was applied to rest fMRI. Time series data were standardized, and a de-spiking approach was applied to correct extreme values for connectivity. Specifically, time points exceeding a threshold of 3 standard deviations were replaced with the average value across all time points for each vertex (Costabel & Müller‐Petke, 2014). GLM analysis was performed using OLS as the noise model, and residual signals were extracted after regressing out nuisance variables and, in the case of task fMRI, task-related activity. The residual time series were then concatenated across runs for each participant.

The residual time series were parcellated into 379 regions. Functional connectivity between regions of interest (ROIs) was computed using Pearson's *r*, followed by Fisher's z-transformation of the correlation matrices. These z-transformed matrices were flattened into 1D arrays for subsequent analysis.

##### Functional MRI (fMRI) Tasks

Emotional Nback task combines working memory demands with emotional processing, using stimuli that include emotional faces (positive, negative, and neutral) and places. Participants perform two conditions: a 0-back task, where they identify whether an image matches one shown at the beginning of the block, and a 2-back task, requiring them to determine whether an image matches one presented two trials earlier. This design allows the task to probe neural activity related to working memory, emotion processing, and facial recognition (Hagler et al., 2019; Jernigan et al., 2023).

The Monetary Incentive Delay (MID) task was designed to investigate reward processing. In this task, participants responded to visual stimuli, with their outcomes determined by their reaction time and the specific condition. Depending on the trial, they could win or lose different amounts of money or have no monetary outcome (Neutral). At the end of each trial, feedback indicated whether they won money (Positive Reward Feedback), missed out on winning (Negative Reward Feedback), avoided a loss (Positive Punishment Feedback), or incurred a loss (Negative Punishment Feedback) (Jernigan et al., 2023; Kennedy et al., 2022).

The Stop Signal Task (SST) task was designed to study inhibitory control through fMRI by requiring participants to suppress or interrupt their motor response to a ‘Go’ stimulus upon seeing a ‘Stop’ signal. This task probes the neural mechanisms underlying response inhibition and motor control(Jernigan et al., 2023; Kennedy et al., 2022).

Using the ABCC, we computed 10 contrasts (see Supplementary figure 1) for each of the fMRI tasks parcellated with Glasser (Glasser et al., 2016) cortical and FreeSurfer(Fischl et al., 2002) subcortical atlases. The contrasts for the Nback task were derived from the design matrix, which represents various task conditions such as stimulus types (places and faces), emotional valences of faces (positive, negative, neutral), and memory loads (two-back and zero-back). These contrasts were constructed to isolate specific effects, including responses to individual stimulus categories (e.g., place or face), combinations of conditions (e.g., emotional faces), and direct comparisons between conditions (e.g., face versus place or positive versus neutral faces). They also allow for the examination of memory load effects by comparing two-back and zero-back conditions. The contrasts for the MID task were derived from the design matrix, which encodes conditions reflecting varying reward and punishment magnitudes (large and small), as well as neutral trials and outcome events (hits and misses). These contrasts were constructed to isolate the neural responses to rewarding versus neutral conditions (e.g., large and small rewards compared to neutral) and similarly for punishment conditions. In addition, contrasts directly compare differences in magnitude (e.g., large versus small rewards or losses) and distinguish between successful versus unsuccessful outcome processing by contrasting hits with misses. The contrasts for the SST task encoded specific task conditions such as stimulus and response type. These contrasts were constructed to isolate neural responses associated with each condition and to directly compare performance differences—such as contrasting CorrectStop with CorrectGo to assess inhibition success, and contrasting IncorrectStop or IncorrectGo with CorrectGo to examine error-related effects. In addition, composite contrasts were created to compare stop versus go trials (e.g., averaging CorrectStop and IncorrectStop relative to CorrectGo) and to differentiate between successful and failed inhibition (e.g., CorrectStop versus IncorrectStop).

The task contrasts available in ABCD tabulated data encompass nine contrasts from Nback, ten from MID and seven from SST parcellated with Destrieux (Destrieux et al., 2010) cortical and FreeSurfer (Fischl et al., 2002) subcortical atlases (see Supplementary figure 1).

##### fMRI Functional Connectivity (FC)

In addition to the region-to-region FC during rest and the three tasks in ABCC, we concatenated the residuals from all available tasks to calculate a multitask FC (Cole et al., 2021) and all available task residuals plus the rest to obtain general FC (Elliott et al., 2019). The rs-fMRI sets of features in ABCD included temporal variance measurements across 333 cortical (Gordon et al., 2016) and 19 subcortical regions. These values represent the variability over time within each parcellated brain region, capturing the amplitude of low-frequency oscillations (Long et al., 2023) ; subcortical-to-network FC (247 features) including the average correlation between each of the 19 subcortical regions and the 13 major cortical networks; and cortical FC (91 features) included the mean correlation values calculated both within and between large-scale cortical networks (Jernigan et al., 2023) (see Supplementary figure 1).

##### Structure MRI (sMRI) and Diffusion Tensor Imaging (DTI)

The ABCD collection includes multiple measures extracted from T1-weighted and T2-weighted 3D structural images using FreeSurfer (Fischl et al., 2002). Cortical and subcortical regions of interest were calculated with Destrieux (Destrieux et al., 2010) and FreeSurfer (Fischl et al., 2002) parcellations respectively. We used FreeSurfer's (Fischl et al., 2002) aparc.stats and aseg.stats files to extract three distinct sets of features within ABCC: cortical thickness, surface area, and FreeSurfer summations. The FreeSurfer summations included five features from FreeSurfer (Fischl et al., 2002): estimated intracranial volume, total cortical gray matter volume, total cortical white matter volume, total subcortical gray matter volume, and the ratio of brain segmentation volume to estimated intracranial volume (Tetereva et al., 2022). We also used 12 other structural sets of features from ABCD tabulated data, including cortical volume, sulcal depth, white-matter averaged intensity, grey-matter averaged intensity, T1 normalized intensity, T1 subcortical averaged intensity, T1 subcortical volume, T1 summations, T2 normalized intensity, T2 subcortical averaged intensity, T2 subcortical volume and T2 summations (Jernigan et al., 2023). Note that T1 summations included average T1 intensity white matter surface, cortical gray matter total area, subcortical gray volume, difference between T1 white and gray matter, cortical gray matter mean thickness, mean sulcal depth, average T1 intensity of cerebellum, average T1 intensity gray matter surface, cortical gray matter total volume. T2 summations included average T2 intensity of cerebellum, average T2 intensity gray matter surface, average T2 intensity white matter surface, difference between T2 white and gray matter.

A high angular resolution diffusion imaging (HARDI) scan was done to segment white matter tracts and measure diffusion parameters. We used fractional anisotropy (FA) of major white matter tracts labelled by AtlasTrack (Hagler Jr et al., 2009). FA quantifies the degree of anisotropy, or directionality, of water diffusion, providing insights into the structural integrity and organization of white matter tracts (Alexander et al., 2007). The processing steps are explained in Hagler and colleagues (Hagler et al., 2019).

Supplementary Figure 1 details all 81 sets of features and their combination in the stacking layer.

#### Lytle and Colleagues’ Pre-processing Pipeline

The dataset was preprocessed using fMRIPrep within a Singularity container. Anatomical preprocessing leveraged pre-computed FreeSurfer (Fischl et al., 2002) derivatives, and functional data underwent standard corrections, including motion correction, normalization to standard space. Outputs were generated in CIFTI format for surface-based analyses. Slice-timing correction was ignored. Next, we applied the first level analysis pipeline explained above to extract task contrasts, task-state and multitask FC. We removed the initial non-steady-state TRs from the time-series, the confounds and the event data based fMRIPrep confounds table. We used FreeSurfer's (Fischl et al., 2002) aparc.stats and aseg.stats files to extract four distinct sets of features: cortical thickness, surface area, subcortical volume and FreeSurfer (Fischl et al., 2002) summations. Supplementary Figure 2 details all 13 sets of features and their combination in the stacking layer.

### Cognitive Prediction Models

**Cross Validation.** In both modelling layers, we applied a nested leave-one-site-out cross-validation scheme: 21 sites (folds) served as test sets iteratively. This process was repeated until every site had been used as the test set once. Within each fold, we standardized the training set features across participants, then applied training statistics (including mean and standard deviation) to normalize the test set before training the predictive model. Within each training set, we performed an additional five-fold cross-validation to optimize the hyperparameters of the predictive models with the negative mean squared error (NMSE) as the evaluation metric. This approach ensures robust and generalizable model evaluation by reducing site-specific biases, preventing data leakage, and providing a realistic estimate of performance on unseen sites by hyperparameter optimization within the training set (Cawley & Talbot, 2010).

In our approach, we first built single models by using Elastic Net regression to predict cognitive-functioning scores from each of the 81 sets of features. For FC data, Partial Least Squares (PLS) was applied as a dimensionality reduction step prior to feeding the resulting components into the Elastic Net. Once these models were trained on the outer-fold training sets (with inner cross-validation for tuning), we computed predicted values from each model. These predictions—generated exclusively from the outer-fold training sets to avoid data leakage—were then used as inputs (one per single model) for training the stacked models. We combined them into 15 different stacking configurations (e.g., stacked all, task contrasts, etc.) and trained a Random Forest model to predict cognitive abilities from each stacked model in the outer-fold training sets (using the same inner-fold cross-validation as single models). For example, to build a stacked model combining task contrasts for ABCD, we first developed single models for each of the contrast-related sets of features (each including 379 or 148 features), and then used the predicted values from these models as individual features in the stacked task contrasts model. This approach, often called late fusion, contrasts with methods that simply concatenate raw features (referred to as early fusion or flat model), and it enables the use of different machine learning algorithms across the various feature sets (Bokade et al., 2021).

Partial Least Squares Regression (PLS) is a dimensionality reduction technique that identifies latent components to maximize the covariance between predictor variables X and the response variable Y, solving for components t and u such that max⁡ Cov(t,u)=wTX and t = Xw, u=Yq, where w and q are weights for X and Y, respectively (Mateos-Aparicio, 2011). To optimize the number of components, we performed a grid search with five-fold cross-validation, considering all components for sets with fewer than 30 features and capping the number at 30 for larger sets of features. The optimal model was selected based on the negative mean squared error (NMSE) and was used to transform the feature space and generate predictions, reducing dimensionality while retaining predictive information (Mateos-Aparicio, 2011).

Elastic Net regression, a regularization technique combining penalties from both Ridge (L2) and Lasso (L1) regression, to balance complexity and prediction accuracy while preventing overfitting. Elastic Net optimizes the following loss function:

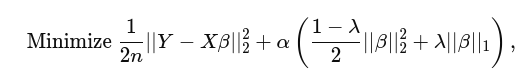

Where ∣∣⋅∣∣1 and ∣∣⋅∣∣2 are the L1 and L2 norms, α controls the overall strength of regularization, and λ balances L1 and L2 contributions (Pedregosa et al., 2024). A grid search was performed to optimize the hyperparameters α (range: 100 to 102) and L1-ratio ([0,1]), with 5-fold cross-validation. The model was evaluated using the negative mean squared error (NMSE) as the scoring metric. The best model was used to predict training and test outcomes. This Elastic Net approach offers a robust solution for handling multicollinearity in predictors while achieving sparse solutions, useful in high-dimensional settings (Zou & Hastie, 2005).

Random Forest regression is an ensemble learning method that aggregates predictions from multiple decision trees to improve predictive performance and reduce overfitting. Its ability to handle non-linear relationships and robustly deal with missing data made it suitable for the stacking layer. Each tree in the forest is trained on a bootstrapped sample of the data, and at each split, a random subset of features is considered, introducing randomness and diversity (Breiman, 2001). The hyperparameters used include the number of decision trees in the forest, set to 1000, the maximum depth of each tree, searched over a range of 1 to 10 to control model complexity, and the number of features considered at each split, with options including the total number of features (None), the square root of the feature count (sqrt), and the logarithm base 2 of the feature count (log2).

***Missingness.*** To address the varying missingness between sets of features, we used the opportunist approach in the stacked layer (Pat et al., 2022). Roughly, we doubled the 81 predicted values, creating two copies of the feature set. Missing values in one copy were replaced with a large positive number (e.g., 1000) and in the other with a large negative number (e.g., -1000). This allowed us to retain participants with at least one valid feature while providing the model with additional information about missingness.

**Evaluation.** Correlation (*r*), coefficient of determination (R²), mean absolute error (MAE), and root mean squared error (RMSE) were reported as measures of predictive performance across all folds for internal-validation-set and for whole external-validation-set, as well as separately for ADHD and non-ADHD subgroups.

We then used bootstrapping to compare model performance between ADHD and non-ADHD groups by resampling predicted and true values 2,000 times. For each bootstrap iteration, mean absolute error (MAE), root mean square error (RMSE), R², and Pearson correlation (PR) are computed for both groups. The differences between these metrics are used to construct 95% confidence intervals, determining statistical significance. We also compared the distributions of observed and predicted cognitive scores between ADHD and non-ADHD groups with a bootstrap procedure (10,000 resamples) to estimate confidence intervals for the mean difference. If the 95% confidence interval did not include zero, the difference was considered statistically significant. Hedges' g is a standardized effect size metric that accounts for sample size, providing a reliable comparison of groups with unequal sample size (National Institute of Standards and Technology (NIST), n.d.).

**Residualization Sensitivity Analysis.** To evaluate the potential influence of demographic, socioeconomic, and medication-related factors on predictive performance, we conducted a post hoc residualization sensitivity analysis. For each model, both predicted and observed cognitive scores were separately residualized using linear regression with parental education, household income, biological sex, and medication exposure as predictors.

Medication exposure was derived from the *su_y_plus* table and classified according to the 14 first-level categories of the Anatomical Therapeutic Chemical (ATC) system (see below for the number of participants in each category). Binary indicator variables (0/1) were then created for each ATC category and entered as predictors in the regression models (Supplementary Table 10).

For each analysis, participants with complete data on the outcome and relevant covariates were included. Residuals from the regressions were treated as covariate-adjusted predicted and observed cognitive scores, and Pearson's r was recalculated using the same paired observations. Analyses were performed separately for each covariate set and for each ADHD status. This procedure was intended as a sensitivity analysis to assess the robustness of model performance and was not incorporated into model training. Full results are presented in Supplementary Table 2.

Full results of the residualization analyses for the Stacked All model are presented in Supplementary Table 2. Overall, residualizing parental education and household income modestly attenuated model performance, whereas residualizing sex and medication exposure had little effect. Importantly, any attenuation was consistent across ADHD status. For example, in the Stacked All model, the predictive correlation (r) in the non-ADHD group decreased from .57 to .50 after adjusting for parental education and to .47 after adjusting for household income, but remained essentially unchanged after adjusting for sex (r = .57) or medication exposure (r = .57). Similar patterns were observed across ADHD Tiers 1-4. These findings suggest that socioeconomic factors contribute modestly to the predictive signal, whereas sex and medication exposure do not appear to explain model performance. Critically, the magnitude of attenuation was comparable across ADHD and non-ADHD groups, indicating that these covariates are unlikely to account for any differences in predictive performance across ADHD status.

**Feature Importance**. In the single layer, Elastic Net coefficients indicated the contribution of each feature within a single set of features. For FC, we calculated the contribution of each feature to the prediction of the target variable by both the weight of each feature in the latent components and the amount of variance explained by each PLS component. In stacked layer two, feature importance was analyzed using SHapley Additive exPlanations (SHAP) values. SHAP quantifies the contribution of individual features by averaging the changes in model predictions when each feature is included versus excluded across all possible feature subsets (Bifarin, 2023; Lundberg & Lee, 2017).

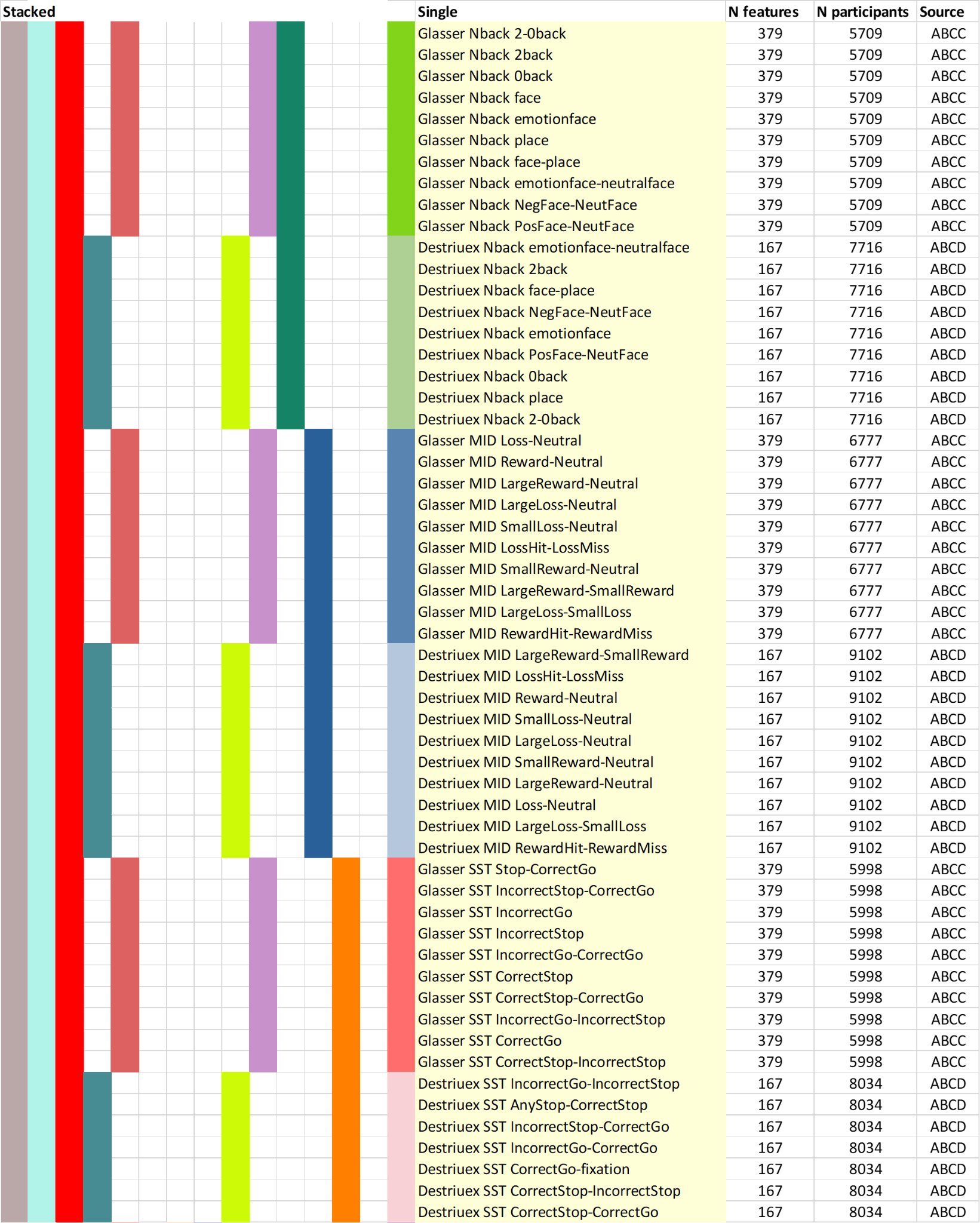

###### Supplementary Figure 1 Single and stacked sets of features in the internal-validation set.

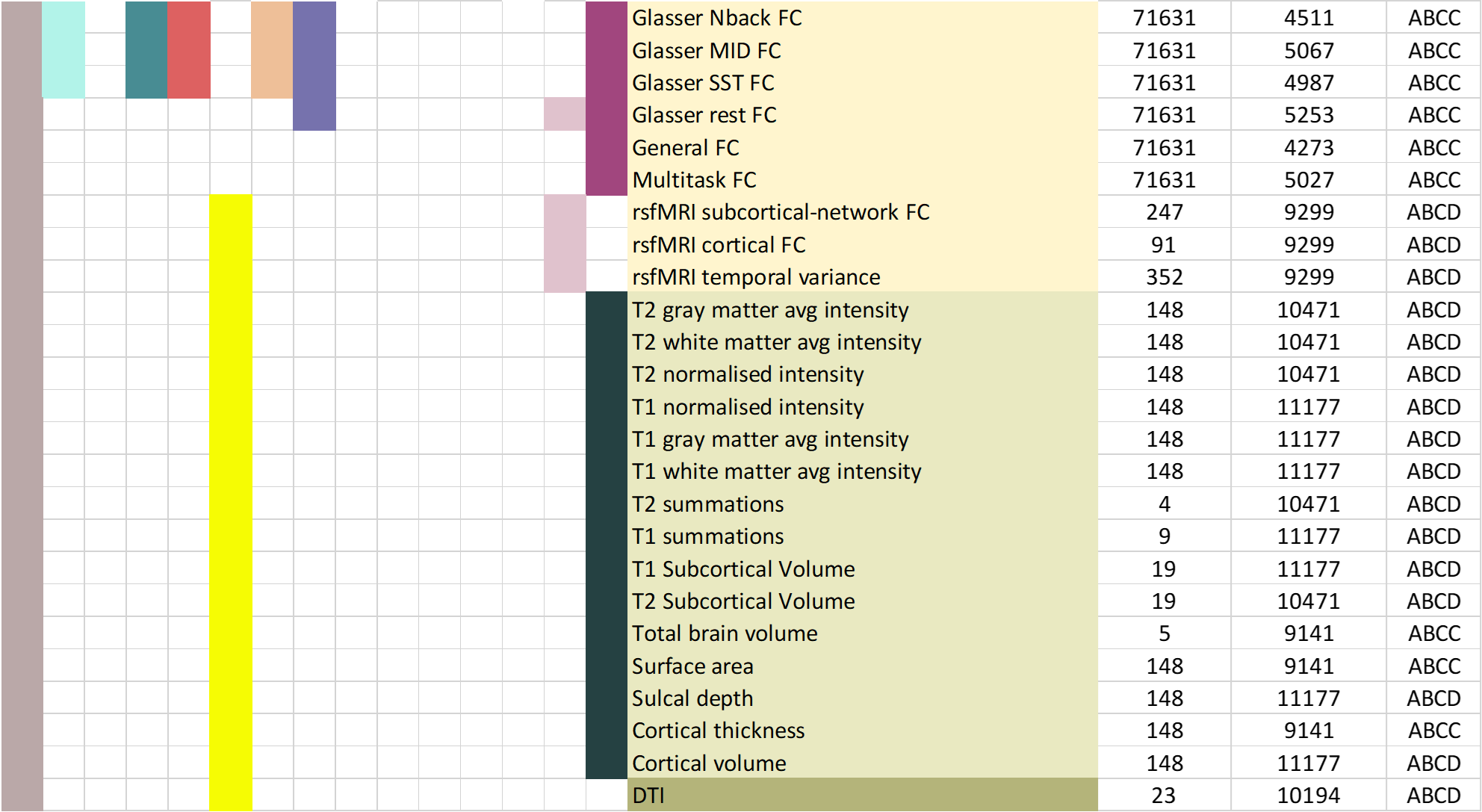

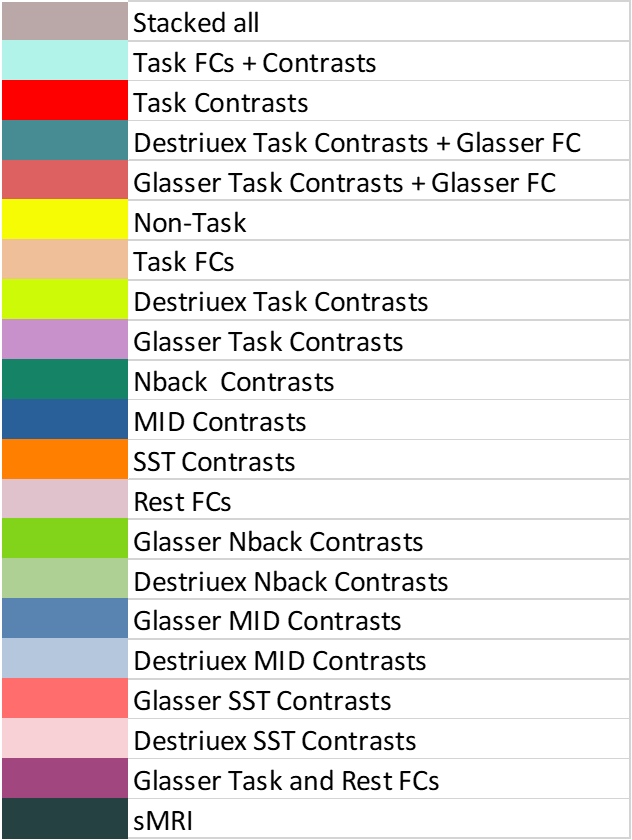

###### Supplementary Figure 2 Single and stacked sets of features in the external-validation set.

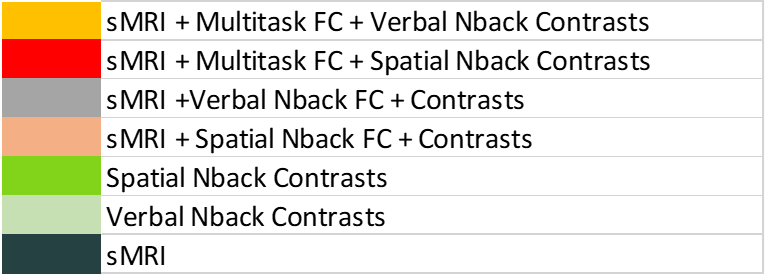

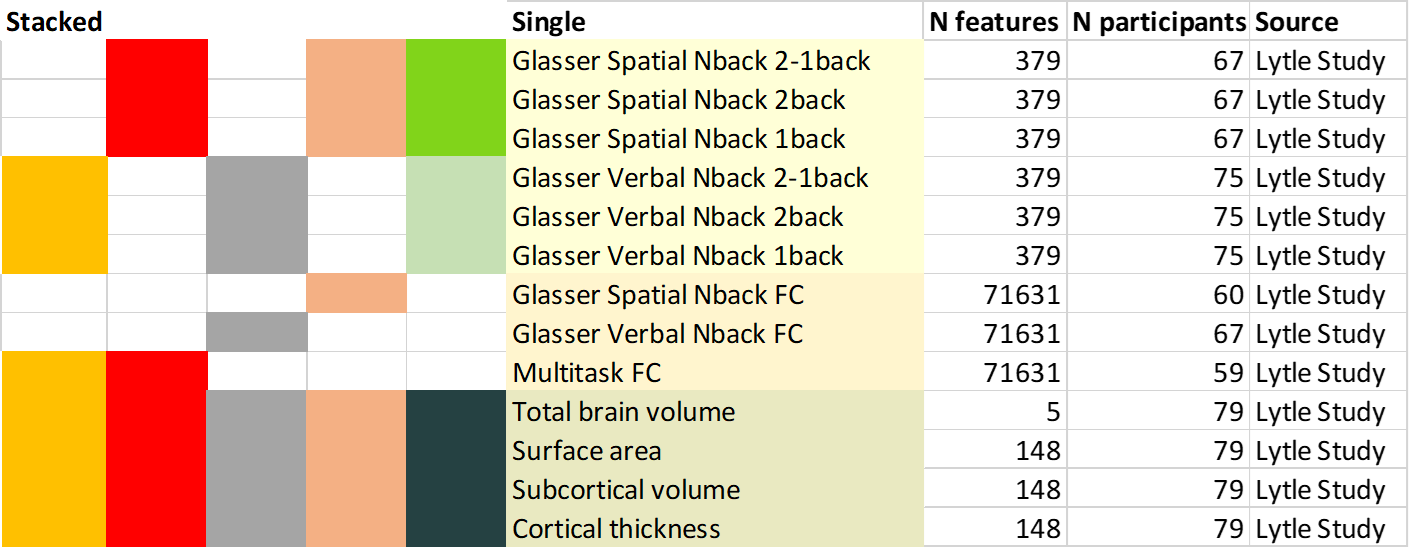
